# Bridging the scales: leveraging personalized disease models and deep phenotyping to dissect cognitive impairment in schizophrenia

**DOI:** 10.1101/2025.02.25.25322779

**Authors:** Florian J. Raabe, David Popovic, Clara Vetter, Genc Hasanaj, Berkhan Karslı, Laura E. Fischer, Emanuel Boudriot, Valeria Almeida, Allesia Atella, Tim J. Schäfer, Miriam Gagliardi, Lucia Trastulla, Vladislav Yakimov, Lukas Roell, Joanna Moussiopoulou, Lenka Krčmář, Sabrina Galinski, Irina Papazova, CDP Working Group, Oliver Pogarell, Alkomiet Hasan, Eva C. Schulte, Andrea Schmitt, Elias Wagner, Anna Levina, Moritz J. Rossner, Nikolaos Koutsouleris, Sergi Papiol, Peter Falkai, Daniel Keeser, Michael J. Ziller

**Affiliations:** Max Planck Institute of Psychiatry, 80804 Munich, Germany; Department of Psychiatry and Psychotherapy, University Hospital, LMU Munich, 80336 Munich, Germany; NeuroImaging Core Unit Munich (NICUM), University Hospital, LMU Munich, 80336 Munich, Germany; Evidence-based psychiatry and psychotherapy, Faculty of Medicine, University of Augsburg, 86156 Augsburg, Germany; Department of Psychiatry, University of Münster, 48149 Münster, Germany; Center for Soft Nanoscience, University of Münster, 48149 Münster, Germany; University of Tübingen, Tübingen, Germany; Max Planck Institute for Biological Cybernetics, Tübingen, Germany; International Max Planck Research School for Translational Psychiatry (IMPRS-TP), 80804 Munich, Germany; Systasy Bioscience GmbH, 81669 Munich, Germany; Department of Psychiatry, Psychotherapy, and Psychosomatics, Medical Faculty, University of Augsburg, 86156 Augsburg, Germany; German Center for Mental Health (DZPG), partner site Munich-Augsburg, Germany; Institute of Human Genetics, University Hospital, Faculty of Medicine, University of Bonn, 53127 Bonn, Germany; Department of Psychiatry and Psychotherapy, University Hospital, Faculty of Medicine, University of Bonn, 53127 Bonn, Germany; Institute of Psychiatric Phenomics and Genomics, University Hospital, LMU Munich, 80336 Munich, Germany; Laboratory of Neurosciences (LIM-27), Institute of Psychiatry, University of São Paulo (USP), São Paulo-SP 05403-903, Brazil; Institute of Psychiatry, Psychology and Neuroscience, King’s College London, London, United Kingdom; Munich Center for Neurosciences (MCN), LMU Munich, 82152 Planegg-Martinsried, Germany

## Abstract

Schizophrenia (SCZ) is a highly heritable brain disorder marked by a wide range of changes throughout the central nervous system. These changes include alterations at the molecular and cellular levels, suggesting significant disruptions in synapse function, as well as modifications in brain structure and activity. However, it remains unclear, how changes in molecular synapse biology translate into neurophysiological and ultimately behavioral consequences across scales. Here, we narrow this translational gap in contemporary biological psychiatry by establishing a generalizable framework to bridge the scales and pinpoint biological mechanisms underlying individual psychiatric symptoms. We show that genetically driven changes in neuronal gene expression and a resulting reduction in excitatory synaptic density *in vitro* are linked to alterations of brain structure, electrophysiology and ultimately cognitive function *in vivo*.

These results provide a direct connection between the molecular origins of synapse reduction in SCZ and its neurobiological and phenotypic consequences on the individual patient level, paving the way to develop new mechanism informed treatment options.

## Introduction

Schizophrenia (SCZ) is a severe mental illness with a lifetime prevalence of 1% of the population and a major cause of global disease burden^1^. Clinical presentation of SCZ is highly heterogenous, with core symptoms affecting three major domains, including positive symptoms (e.g. hallucinations, paranoia), negative symptoms (e.g. anhedonia, social withdrawal) and cognitive symptoms (e.g. impaired working memory and attention deficits)^2^. Particularly the latter impairments vary substantially in severity across individuals and significantly impact overall functioning, quality of life, and occupational success^2–4^. While current antipsychotics can effectively improve positive symptoms, addressing cognitive impairments in SCZ remains a critical and unmet medical need^2^.

At present, the biological causes and neurobiological mechanisms underlying SCZ and in particular cognitive impairment remain elusive. One major hypothesis in the field proposes that SCZ ultimately originates from deficits in synaptic function, arising from a variety of molecular mechanisms^5^. In its current form, this hypothesis suggests a multi-hit model, where genetic and environmental risk factors (e.g. maternal infection, drug abuse etc.) render synapses vulnerable to excessive microglia mediated elimination over the course of late neurodevelopment disrupting micro- and macro-connectivity of brain circuits^5^.

This hypothesis is supported by several lines of evidence, including complementary findings from genetic and neuroimaging studies^6,7^. On the genetic level, genome wide association studies (GWAS) have identified hundreds of SCZ associated loci that are strongly enriched for immune, neurodevelopmental and synapse biology related genes^8^. Moreover, functional follow up studies in human *in vitro* and *in vivo* animal models imply genetically and/or stress induced increased microglia driven synaptic pruning as one mechanism contributing to a reduction in synapse number in SCZ^5^. However, independent of microglia and pruning related genetic risk factors, the same GWA studies identified dozens of additional genes directly involved in synaptic function and neurotransmission. These largely remain of unknown relevance for disease etiology at present^8^.

On the neurophysiological level, meta-analyses of neuroimaging data consistently demonstrate structural gray matter volume (GMV) reductions in individuals with schizophrenia compared to healthy controls^9–13^. Results from post-mortem brain and animal studies indicate, that these changes originate predominantly from a reduction in dendritic volume and synaptic density, while neuronal and glia soma number remained unaltered^5,14,15^.

These widespread GMV alterations are observed across multiple brain regions, reflecting the distributed nature of neuropathological changes in the disorder. Among the most consistently implicated regions are the anterior cingulate cortex, bilateral insular cortices, the dorsolateral prefrontal cortex (DLPFC) and temporal lobe regions, which are critical areas for executive functions and working memory. For instance, reductions in DLPFC GMV have been specifically linked to cognitive deficits characteristic of schizophrenia, such as impairments in planning, attention, and cognitive flexibility^9,13^.

Electrophysiological studies using electroencephalography (EEG) in individuals with schizophrenia consistently demonstrate alterations in neural oscillations, particularly within the theta and gamma frequency bands^16,17^. These aberrant oscillatory patterns, reflect disrupted neural synchrony and are implicated in the cognitive deficits characteristic of SCZ, including impairments in working memory, attention, and executive function. Therefore, EEG-derived measures of theta and gamma activity represent a promising avenue for understanding the macroscale neurophysiological mechanisms contributing to cognitive dysfunction in schizophrenia.

However, at present these observations remain largely disconnected. Both the molecular underpinnings as well as the neurobiological and behavioral consequences of these findings on the genetic, post-mortem and brain structural level remain elusive.

This missing link between molecular and cellular level changes leading to a reduction in synaptic density, SCZ specific macro-circuit and neurophysiological alterations as well as ultimately behavioral phenotypes represents a major translational gap in contemporary mental health research. Jointly, this lack of insight severely impedes progress in the identification of molecular and neurobiological mechanisms contributing to specific symptom domains. Moreover, these missing links hinder the identification of disease and symptom relevant drug targets and the development of new treatment strategies targeting the consequence of synapse reduction. On the clinical level, narrowing this gap can pave the way towards the identification of brain regions or circuits where synaptic loss contributes to specific SCZ associated neurophysiological and behavioral changes. Selectively targeting specific regions or circuits in the brain that contribute to individual symptom classes such as cognitive impairment, holds great potential to improve notoriously hard-to-treat neurocognitive performance.

Previous strategies to narrow this gap have relied on computational approaches and biophysical neural mass models of micro- and macro-circuits to simulate EEG or resting state functional magnetic resonance imaging (MRI) data^18,19^. By systematically evaluating circuit connectivity parameters (e.g. synaptic strength) *in silico*, these studies showed that alterations in excitatory synaptic gain^18^ or changes in excitation to inhibition ratio in cortical circuit models produced EEG or resting state fMRI traces consistent with empirical data from SCZ patients *in vivo*^19^. However, at present these results remain on the level of statistical inference, fitting model parameters to empirical macro-scale observations, thereby lacking an empirical molecular and cellular basis.

More recently, personalized disease models based on patient-derived induced pluripotent stem cells (iPSCs) emerged as another promising bridging technology. These in models provide the opportunity to dissect disease-relevant molecular mechanisms in patient-derived cells *in vitro*^20–22^. Conceptually, iPSCs capture the entire polygenetic risk profile of the original donor, enabling the assessment of their joint molecular and cellular effects in disease-relevant brain cell types^20^. Consequently, iPSCs offer a pivotal tool for understanding the previously inaccessible consequences of the complex genetic architecture of SCZ. Most importantly, they unlock the possibility to connect molecular and cellular level alterations *on the micro scale* to macroscale disease manifestations and clinical intermediate phenotypes of the same patients^23^. We recently utilized this technology to identify one genetically driven mechanisms causing a reduction of synaptic density in iPSC derived neurons from SCZ patients, independent of microglial pruning^24^. Instead, reduced synaptic density was driven by altered expression and alternative polyadenylation of many synaptic transcripts, partially mediated by the SCZ specific elevated expression of multiple RNA binding proteins.

In this study, we set out to bridge the translational gap in biological psychiatry and determine how alterations in molecular and cellular synapse biology translate into neurophysiological and ultimately behavioral consequences across scales. To this end, we address three key questions:

1. What are the behavioral consequences of changes in region specific synaptic density changes based on GMV reductions?
2. What are the neurophysiological consequences of altered region specific GMV based synapse density?
3. Do these changes on the structural and neurophysiological level in part originate from the individual level complex genetic liability to SCZ?

To answer these questions, we developed a translational research framework that bridges the scales by integrating deeply phenotyped clinical cohorts^25^ with a corresponding molecularly characterized iPSC cohort^24^, machine learning and biophysical circuit modeling.

## Results

### Linking Structural Brain Alterations to Cognitive Impairment in SCZ

To determine the behavioral consequences of changes in region specific synaptic density, we considered alterations in GMV as approximation. Based on previous evidence indicating structural brain alterations in SCZ^6,10,26^, we hypothesized that region specific changes in GMV might particularly contribute cognitive impairment in SCZ as one critical clinical phenotype. To test this hypothesis, we sought to identify a multimodal signature of cognitive alterations in SCZ and their link to structural brain changes in two deeply phenotyped cohorts (**Fig. 1a** and **1b, Supplementary Table S1)**. Therefore, we first performed multivariate and unsupervised analysis between cognitive and clinical parameters (patient phenotypes) and whole brain GMV structural data from T1 MRI scans of 131 individuals within our discovery cohort C1 from the Multimodal Imaging in Chronic Schizophrenia Study (MIMICSS)^25^. Specifically, we utilized the SPLS algorithm^27^, an iterative, linear, algorithm based on singular value decomposition, which identified two significant signatures, each representing a distinct combination of phenotypic and structural brain patterns.

**Fig. 1:**
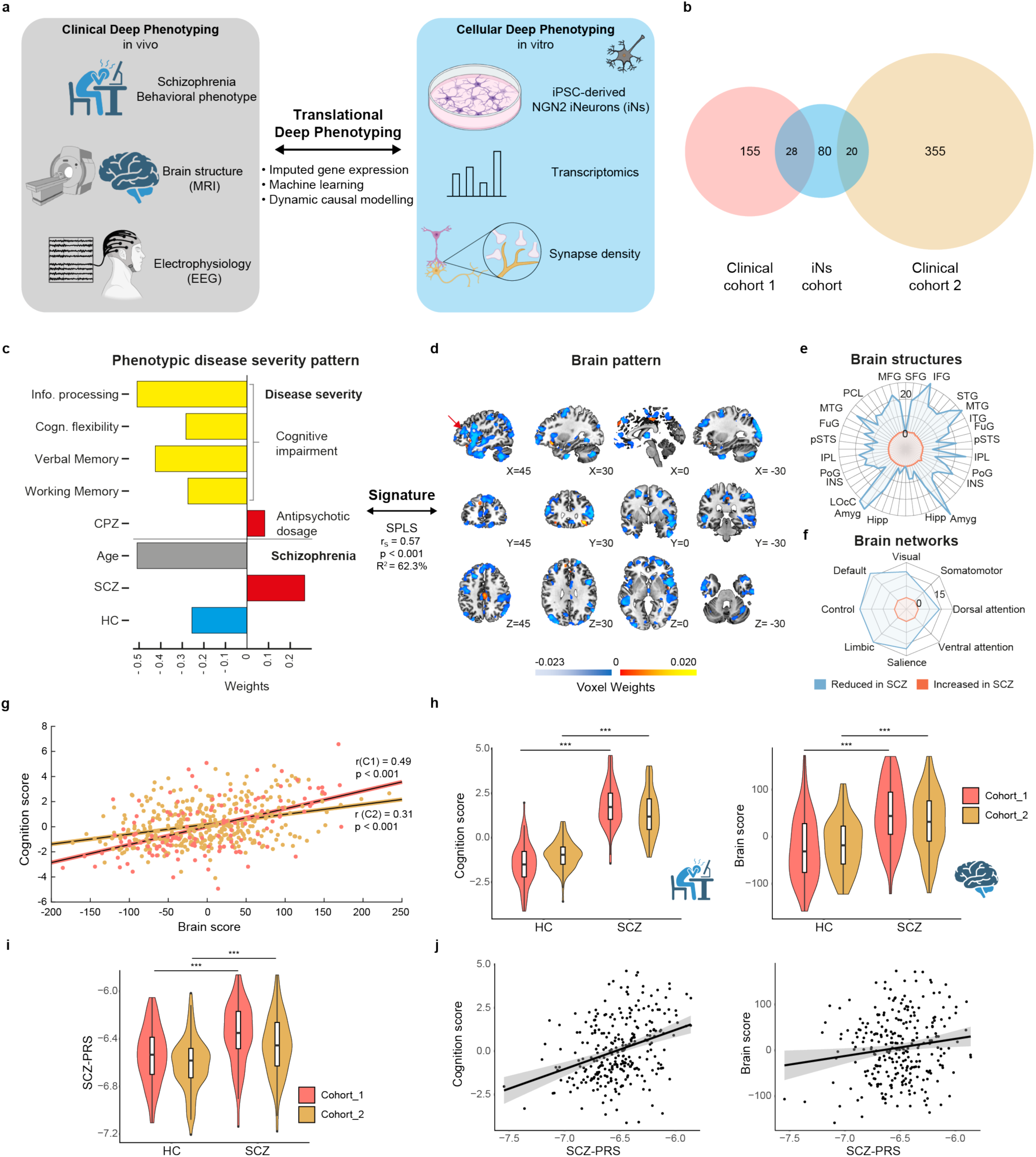
Scheme of Translational Deep Phenotyping to investigate a multivariate phenotype-brain signature of cognitive impairments in schizophrenia. **a,** Schematic overview of the concept of ‘translational deep phenotyping’ based on a deep phenotyped cohort ^25^ with extended clinical characterization, neuropsychological testing, MRI, and EEG, that overlaps with an hiPSC cohort ^24^ to establish links between in vitro and in vivo scales and to bridge the ‘translational gap’ between genetic, molecular, and cellular in vitro phenotypes and the individual intermediate phenotype of cognitive impairment in individuals with schizophrenia (SCZ). **b,** Venn plot illustrating the sizes of the clinical and hiPSC cohorts and their overlap available for analysis. **c,** Barplot visualizing the phenotypic pattern of the *‘schizophrenia neurocognition signature’* captured by Latent Variable 2 (LV2) derived by the multivariate sparse partial least squares (SPLS) analysis. The arrow indicates the Pearson coefficient (r) and associated p-value as well as the coefficient of determination (R^2^) between the cognition pattern and the brain pattern within the signature of LV2 (N = 131, r = 0.57, p < 0.001, t-test, R^2^ = 62.3%, N=131). **d,** Illustration of the corresponding grey matter volume brain pattern of the *‘schizophrenia neurocognition signature’* (LV2). Positive weighting of voxels is indicated in the red and negative weighting in the blue color scale. **e,** Spider plot highlighting the top neuroanatomic regions captured in the LV2 brain pattern, depicting the proportion of positively (red) and negatively (blue) weighted voxels of LV2 in each region according to Brainnetome and Diedrichsen atlas^45,46^. *Abbreviations: Amyg, Amygdala; FuG, Fusiform Gyrus; Hipp, Hippocampus; IFG, Inferior Frontal Gyrus; INS, Insular Gyrus; IPL, Inferior Parietal Lobule; ITG, Inferior Temporal Gyrus; LOcC, Lateral Occipital Cortex; MFG, Middle Frontal Gyrus; MTG, Middle Temporal Gyrus; PoG, Postcentral Gyrus; pSTS, Posterior Superior Temporal Sulcus; STG, Superior Temporal Gyrus*. **f,** Mapping of the LV2 brain signature onto 8 large-scale functional brain networks, derived from the Yeo (cerebrum) and the Buckner atlas (cerebellum)^47,48^, showing the proportion of positively (red) and negatively (blue) weighted voxels of LV2 in each of these brain networks. **g,** Correlation of individual brain and cognition scores from the *‘schizophrenia neurocognition signature’* from patients with SCZ and healthy controls in cohort 1 (C1, black colored, N =119, r =0.49, p <0.0001, t-test) and cohort 2 (C2, grey colored, N =160, r =0.31, p <0.0001, t-test) with linear regression lines. **h,** Left: Group-level comparison of individual cognition scores from the *‘schizophrenia neurocognition signature’* between healthy control individuals (HC, N_C1_=66, N_C2_=84) and SCZ (N_C1_=53 N_C2_=76) individuals in the discovery (C1, red) and replication (C2, orange) cohort. The significance of differences between groups was evaluated using the Wilcoxon-sum rank test (*** = p < 0.001). Right: Group-level comparison of individual brain scores from the *‘schizophrenia neurocognition signature’* between healthy control individuals (HC, N_C1_=66, N_C2_=84) and SCZ (N_C1_=53 N_C2_=76) individuals in the discovery (C1, red) and replication (C2, orange) cohort. The significance of differences between groups was evaluated using the Wilcoxon-sum rank test (*** = p < 0.001). **i,** Group-level comparison of the SCZ polygenic risk score based on the PGC wave 3 weights between healthy control individuals (HC, N_C1_=65, N_C2_=84) and SCZ (N_C1_=52 N_C2_=76) individuals in the discovery (C1, red) and replication (C2, orange) cohort. The significance of differences between groups was evaluated using the Wilcoxon-sum rank test (*** = p < 0.001). **j,** Correlation analysis of SCZ polygenic risk score (SCZ-PRS, x-axis) and cognition (left, y-axis) or brain score (right, y-axis) across both cohorts jointly (N=272). Black lines indicate regression line based on linear models correcting for the first three principal components of the ancestry matrix for the brain score (r=0.16, p-value=0.0131, t-test) and the cognition score (r=0.36, p-value=34e-09, t-test). Grey shading indicates the estimated standard error at each point.

The *‘aging signature’* represented a brain pattern of widespread GMV reduction (Spearman’s *ρ* = 0. 78, *p* < 0.001, *N* = 131; **Extended Data Fig. 1, Supplementary Table S2-3**), in line with the well-known general pattern of grey matter reduction during aging^28,29^.

The ‘*schizophrenia neurocognition signature’* (Spearman’s *ρ* = 0. 57, *p* < 0.001, *N* = 131) linked a clinical pattern of younger (negative weight for age) individuals with SCZ (positive weight) suffering from pronounced cognitive impairments (negative weight) (**Fig. 1c**) to a brain pattern of mainly GMV reductions (**Fig. 1d**) independent of the age effect already captured by the *‘aging signature’* **(Supplementary Table S2-3**). In the cognitive domains, this affected particularly information processing, cognitive flexibility, verbal long-term memory and working memory (**Fig. 1c**), linking them to GMV in the frontal gyrus, temporal gyrus, right insula, amygdala and hippocampus (**Fig. 1e)**. On the macro-ciruit level, the GMV brain pattern mapped predominantly to the default, limbic, Control and Salience networks (**Fig. 1f, Supplementary Table S4-5**).

Thus, the ‘*schizophrenia neurocognition signature’* captures a profile of younger patients with severe cognitive impairments that are associated with pronounced grey matter loss within macro-circuits critical for cognitive function^12,30,31^. For replication of the SPLS ‘*schizophrenia neurocognition signature’* we leveraged 153 individuals from a second, independent translational deep phenotyping cohort (C2, *Clinical Deep Phenotyping Study (CDP),* **Supplementary Table 1**) (**Online Methods**). Thereby, we confirmed the generalizability of the ‘*schizophrenia neurocognition signature’* by correlating their respective brain and phenotype scores, despite slightly different study protocols (**Fig. 1g**).

In summary, these analyses identified a reproducible pattern of GMV reduction in SCZ and associated cognitive-behavioral signature. This signature can be operationalized to score each individual with respect to the individual loading onto the brain-structural and clinical-phenotypic dimensions of the ‘*schizophrenia neurocognition signature’*. Application of this concept enabled us to capture alterations in the high-dimensional clinical and brain structure space for each individual with dimensions that we term the *Cognition* and *Brain* score. These scores are significantly higher in SCZ patients compared to unaffected healthy controls (HC) in the discovery (p-value_BrainScore_= 6.8e-07, p-value_CognitionScore_ = 1.15e-18, Wilcoxon-test) and in replication cohort (p-value_BrainScore_= 8.7e-06, p-value_CognitionScore_ = 1.11e-22, Wilcoxon-test) (**Fig. 1h**), linking cognitive changes to region specific reduction in synapse density based on GMV.

### Molecular basis of alterations in individual patient level brain intermediate phenotypes

To identify the molecular and cellular basis of SCZ associated changes in the *Brain* and *Cognition* scores of the ‘*schizophrenia neurocognition signature’*, we tested the hypothesis whether these scores exhibited a disease associated heritable component. Correlation analysis of both scores with SCZ polygenic risk^8^ (**Fig.1i**), confirmed a significant correlation (p-value_BrainScore_=0.0131, p-value_CognitionScore_ = 2.34e-09, t-test; **Fig.1j**). These results indicate that the *Brain* and *Cognition* scores of the ‘*schizophrenia neurocognition signature’* capture trait specific aspects of the illness, rendering them amenable to further genetic dissection.

Based on these findings and our previous study^24^, we hypothesized that particularly genetically driven alterations in synapse related gene expression pattern contribute to the emergence of this signature. To test this hypothesis, we utilized an established iPSC cohort^24^ (n=80) of SCZ patients and healthy control (HC) individuals from the translational deep phenotyping cohort^25^ (**Fig. 1b**). Previously, we described the *in vitro* differentiation of this iPSC cohort into cortical pyramidal excitatory neurons (iNs), giving rise to a deeply phenotyped personalized disease model library, including whole transcriptome profiles^24^.

Given that the iPSC library comprised only a subset of the clinical cohort, we leveraged the concept of transcriptome imputation^32^, predicting the gene expression pattern in iNs from the individual level genotype using machine learning^33^ (**Extended Data Fig. 2a**). The imputed gene expression levels showed good agreement with measured transcriptomes (median pearson correlation coefficient r= 0.399, **Extended Data Fig. 2b**). Moreover, differential gene expression patterns between SCZ and HC in iNs between predicted and measured log_2_ fold-changes were correlated (r=0.45, p-value< 2.2e-16, F-test) with 75% of nominally significant genes exhibiting concordant direction of effect (**Extended Data Fig. 2c-e**). Thus, these imputed transcriptome profiles capture a significant fraction of the genetic component of personalized disease model gene expression patterns.

We utilized these imputed gene expression profiles to identify the molecular basis of the *Brain* and *Cognition* scores as intermediate phenotypes. Therefore, we predicted the individual levels of both scores from the experimentally identified DEGs in iNs (N=310) using support vector regression (SVR) in a repeated nested cross-validation pipeline, excluding individuals with empirically measured transcriptome profiles. This machine learning campaign revealed a predictive capacity of the imputed transcriptome of r=0.76 (p-value=0.0002, permutation-test) for the phenotype and r=0.39 (p-value=0.0002, permutation-test) for the brain score (red lines **Fig. 2a Supplementary Table S6-7**) with an associated mean AUC*_CognitionScore_* of 0.91 and AUC*_BrainScore_*=0.69 in the discovery cohort C1 (**Fig. 2b**). Application of the trained SVR model to a held-out patient sample with empirically measured rather than imputed transcriptome profiles revealed a similar predictive performance (**Fig. 2a**, and **b**, blue line, cognition score: r=0.77, p-value =0.0002, AUC=0.95; brain score: r=0.31, p-value =0.049, AUC=0.6 permutation-test). Similarly, replication analyses based on a second, independent cohort C2 (CDP) using the SVR model trained on C1 confirmed the generalizability of these results for the *Cognition score* (r=0.17, p-value=0.017, AUC=0.63, permutation-test) and *Brain score* respectively (r=0.23 p-value=0.0038, AUC=0.58, permutation-test) (**Fig. 2a,b** orange lines). Notably, these results were specific to the selected genes feature set as the imputed as well as empirical DEG iN datasets significantly outperformed 100 models trained on randomized sets of genes (**Fig. 2c, Supplementary Table 8**).

**Fig. 2:**
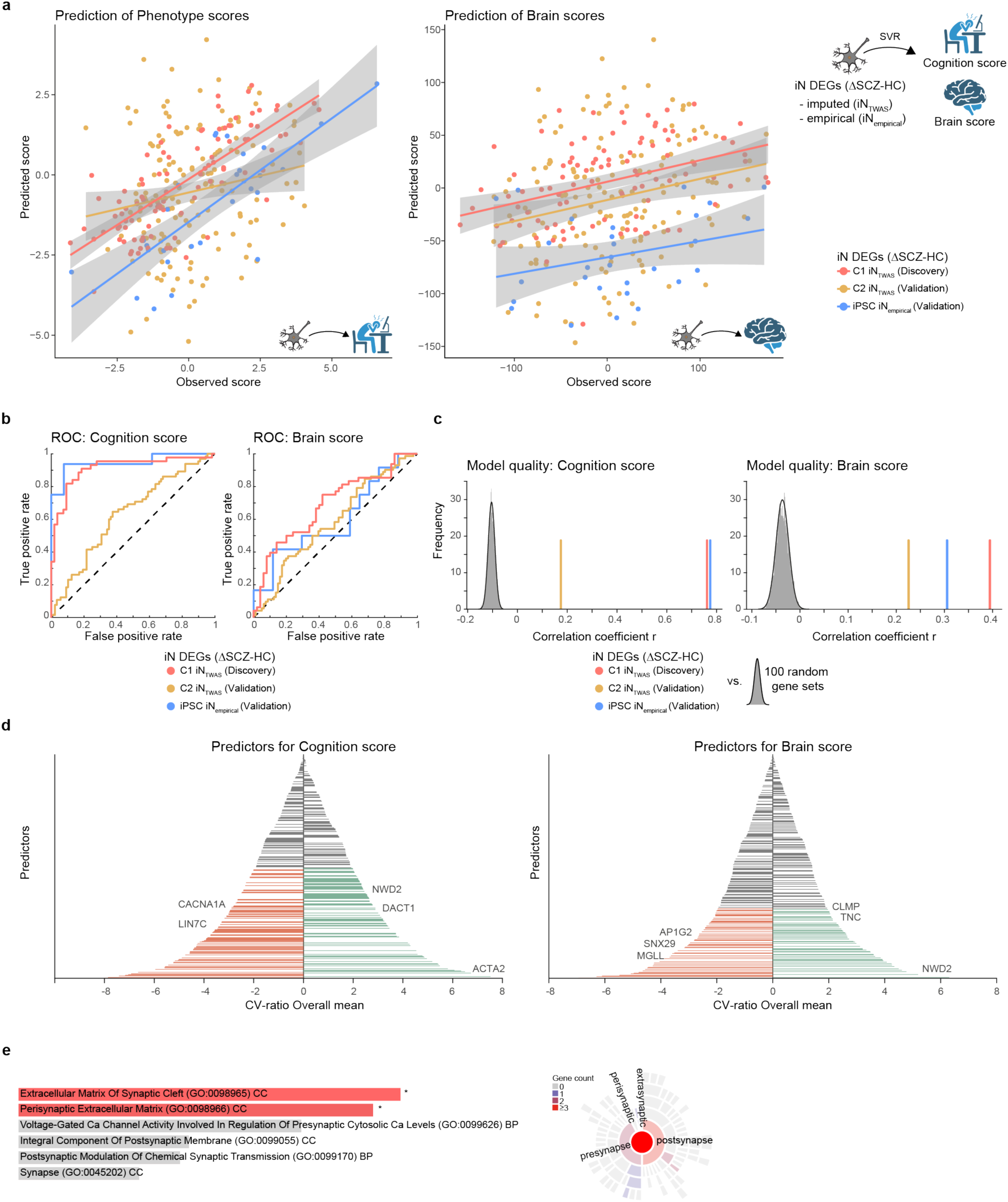
hiPSC-based neuronal transcriptome predicts individual cognition and brain scores. **a,** Correlation between the predicted (y-axis) and observed (x-axis) cognition (left) and brain (right) scores from the *‘schizophrenia neurocognition signature’* LV2. Prediction of the respective scores was performed using support vector regression (SVR) and genotype imputed gene expression levels of 310 genes differential expressed genes (DEGs) in iPSC derived iNs (iN_TWAS_). The latter were previously identified to be differentially expressed in iPSC derived iNs (see **Extended Data Fig. 2c**) and were imputed from genotype in all individuals of the discovery and replication cohort separately. The SVR model was trained on this feature set in the discovery cohort (C1, red) using 5-fold cross validation and subsequently applied to the independent replication cohort (C2, orange) and held-out empirically measured (not-imputed) iN transcriptome data (iPSC-iN, blue). Lines indicate linear regression model for all three cohorts for the cognition (N_C1_=98, r_C1_=0.76, p_C1_=0.0002, N_C2_=153, r_C2_=0.17, p_C2_= 0.018, N_iN_=29, r_iN_=0.77, p_iN_=0.0002, permutation-test) and brain scores (N_C1_=98, r_C1_=0.39, p_C1_=0.0002, N_C2_=153, r_C2_=0.23, p_C2_=0.0038, N_iN_=29, r_iN_=0.31 p_iN_=0.05, permutation-test), with empirically measured having a different offset. **b,** The receiver operating characteristic (ROC) curves and area under the curve (AUC) values describing the SVR model performance for all models in a. for the cognition score (left) in the discovery (C1, red, median AUC=0.89), replication (C2, orange, median AUC=0.57) and empirical replication (iN, blue, median AUC=0.89) cohort as well as the brain score (right) in the discovery (C1, red, median AUC=0.69), replication (C2, orange, AUC=0.57) and empirical replication (iN, blue, AUC=0.56) cohort. **c,** Permutation based evaluation of feature specificity of the SVR model, showing the frequency (y-axis) of the Spearman correlation coefficient (r, x-axis) between the observed and predicted cognition scores (left) or brain scores (right) using the same SVR strategy but based on 100 randomly generated gene feature sets (grey bars) as empirical background distribution. SVR model results from a. are shown as bars for comparison across the three cohorts (C1 – red, C2-orange, iN-blue). **d,** Feature importance plots of the SVR models in a. depicting the ranked cross-validation ratio overall means (CVR-OM, y-axis) of all 310 DEGs gene features (y-axis) in the SVR model to predict the cognition (left) or brain score (right) SVR. Positive CVR-OM indicates that the feature (imputed gene) was predictive of higher cognition or brain scores, whereas negative CVR-OM indicates the opposite. Significant features are magnified and colored, with annotations for selected genes of interest. **e,** Left: Bar plots from gene enrichment analysis based significant CVR-OM genes that predicted brain and cognition score using Enrichr^49,50^ based on SynGo ontology^51^. Enriched pathways sorted by v-value ranking. * indicate pathways that are significantly enriched with p < 0.05. Right: Sun burst plot illustrating locations of synaptic CVR-OM genes. Colour scheme indicates number of genes per term.

Annotation based analysis of the predictive gene features revealed the contribution of numerous genes (**Fig. 2d**) that are significantly enriched for synapse related biological processes (**Fig. 2e**), including several neurotransmission related genes such as the Calcium Voltage-Gated Channel Subunit Alpha1 A (CACNA1A), Dishevelled binding antagonist of beta catenin 1 (DACT1) or Sorting Nexin 29 (SNX29) previously associated with neuronal and cognitive function^34,35^.

Jointly, these findings show that personalized disease model based molecular features are predictive of complex multi-modal cognitive and brain structural phenotypes across patients with SCZ. These results imply alterations in the expression patterns of synaptic genes as one important contributor to cognitive impairment. They support the notion that the observed regional reduction in GMV arises in part from a genetically driven reduction in synapse density, independent of microglial pruning^6^.

### Neurophysiological consequences of altered brain structure as functional basis for cognitive impairment in SCZ

Although these findings shed light on the molecular and brain structural basis of behavior phenotypes in SCZ such as cognitive impairment, they do not elucidate the underlying neurophysiological mechanisms. Therefore, we hypothesized that the molecular and structural changes observed in SCZ are linked to alterations in neuronal and circuit activity, as a potential neurophysiological consequence.

We therefore investigated EEG measurements, where alterations have previously been associated with SCZ and cognitive performance^36^. On the physiological level, EEG patterns represent the aggregated electric signal of the postsynaptic potential of cortical pyramidal neurons^37^. In line with previous findings^38^, EEG power spectrum analysis of 217 individuals with SCZ and 139 HC under eyes-closed resting state conditions revealed a significant increase in theta power and a decrease in Gamma 1 and Gamma 2 power in patients with SCZ compared to unaffected HCs (**Fig. 3a** and **b**).

**Fig. 3:**
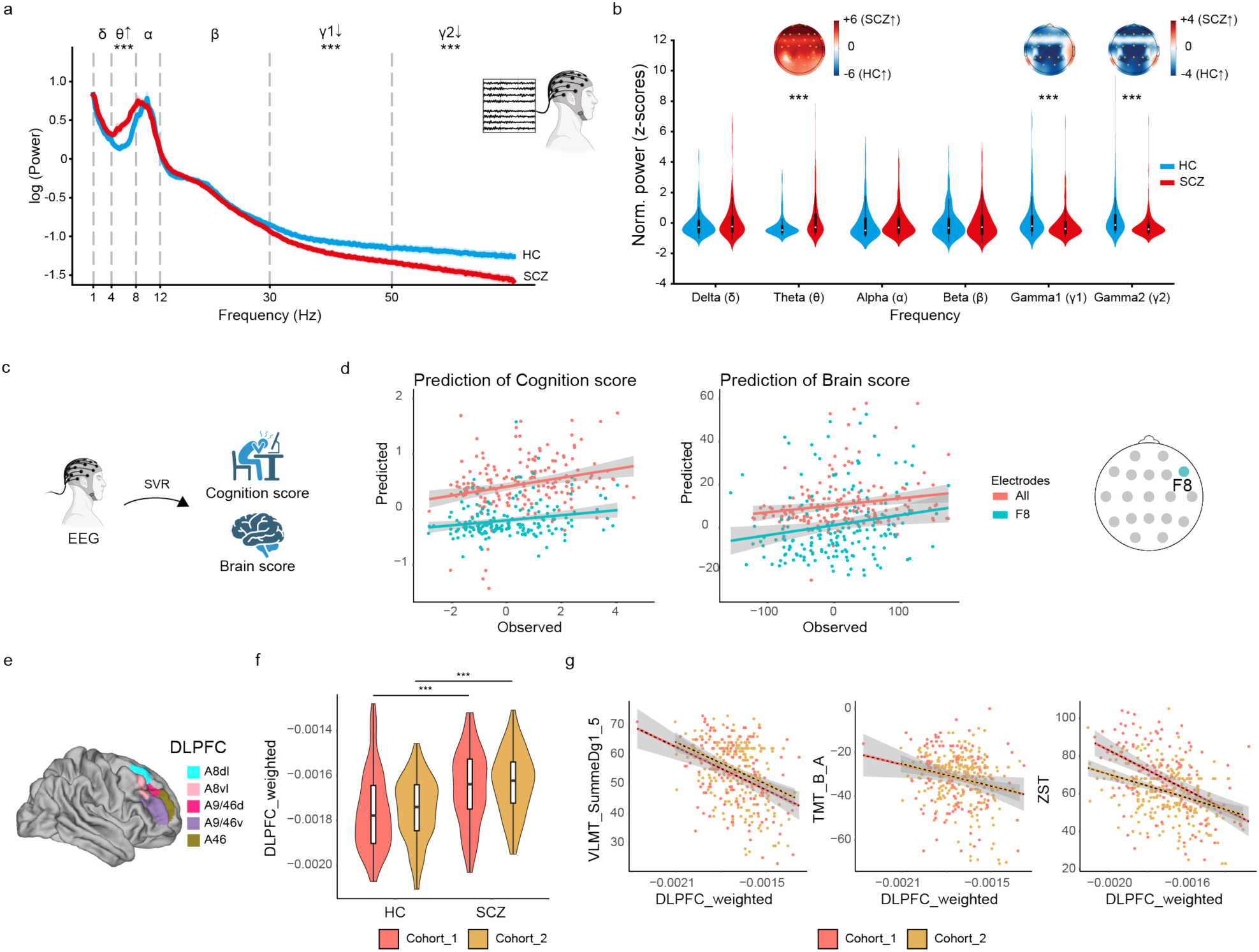
EEG power spectrum alterations in schizophrenia relate to the individual molecular signature. **a,** EEG power spectrum, recorded in resting state with eyes closed from 139 HC (blue line) and 217 patients with SCZ (red line) across analyzed frequency bands (1 Hz to 70 Hz). EEG power spectrum frequency bands are highlighted: delta (δ), theta (θ), alpha (α), beta (β), gamma1 (γ1), and gamma2 (γ2). Arrows denote significant differences in SSD compared to HC by Mann-Whitney-U Test and *** indicating p(FDR) < 0.001. **b,** Violin plots depicting the normalized power (z-scores) distribution for each EEG power spectrum frequency band in 139 HC (blue) and 217 patients with SCZ (red) based on EEG eyes closed recordings. Differences between the groups were evaluated using Mann-Whitney-U Test (*** indicates significant differences at p(FDR) < 0.001). Topoplots above the violin plots illustrate the spatial distribution of EEG power across the scalp, indicating areas with altered power spectrum of heightened (stronger red intensity) or diminished (more bluish tones) activity in SSD compared to HC. **c,** Illustration of the prediction of the individual cognition and brain scores of the *‘schizophrenia neurocognition signature’* LV2 using the individual power of the EEG frequency bands. **d,** Correlation analysis between the predicted (y-axis) and observed (x-axis) cognition (left) and brain scores (middle) of the *’schizophrenia neurocognition signature’* LV2 based on a nested cross-validated support vector regression (SVR) predicting the latter scores from individual level EEG frequency band power in the theta, gamma1 and gamma2 frequency range across all electrodes and the joint C1 and C2 cohorts (red). All depicted predicted values result from prediction where the respective sample was held out in the outer fold of the nested cross-validation. Regression line shows a significant association between predicted and observed cognition (N=191, r=0.2, p-value= 0.0002, permutation-test) or brain score (N=191, r=0.17, p-value= 0.019, permutation-test). In addition, SVR based predictions using only the same features for the F8 electrode (orange) are shown for the cognition (left, N=191, r=0.36, p-value=0.0002, permutation-test) or brain score (middle, N=191, r=0.28, p-value=0.0002, permutation-test). Topo plot on the right indicates the anatomical location of the F8 electrode as the top ranking predictive feature with highest sum of cross-validation ration overall mean (CVR-OM) in the cognition and brain score SVR analysis. **e,** Schematic of anatomical regions considered for the right DLPFC. **f,** Distribution of GMV averaged across all voxels within the right DLPFC weighted by the voxel level feature weight in the SPLS for each individual separated by cohort for individuals with SSD and healthy donors (Ctrl). Distributions differ significantly between the two groups in each cohort (Cohort 1: p-value=0.0002414, Cohort 2: p-value= 1.497e-09, wilcoxon-test). **g,** Correlation of weighted GMV in the right DLPFC (x-axis) and cognitive-testing parameters (y-axis, VLMT_SummeDg1_5-Verbal Learning and Memory Test summary of scores, TMT_B_A-Trail Making Test results of part A an B, ZST-Number Symbol Test) in for each individual and both cohorts. Shown correlations are all significant in each cohort (VLMT_SummeDg1_5: r_Cohort1_=0.3531289, p-value_Cohort1_=5.34e-05, r_Cohort2_=0.3708099, p-value_Cohort2_=7.33e-10; TMT_B_A: : r_Cohort1_=0.3010648, p-value_Cohort1_=0.000645, r_Cohort2_=0.2104044, p-value_Cohort2_=0.000654; ZST: : r_Cohort1_=0.3010648, p-value_Cohort1_=0.000645, r_Cohort2_=0.3676955, p-value_Cohort2_=1.04e-09, t-Test)

To test the hypothesis that these changes in neuronal activity represent a potential consequence of changes in GMV, we evaluated the link between the *Brain score* of the ‘*schizophrenia neurocognition signature’* and the brain’s electrophysiological resting-state activity EEG pattern (**Fig. 3c**). SVR based prediction of the individual *Brain score* from the matching patients’ resting-state EEG power spectrum revealed a significant association between the two layers (**Fig. 3d**, r=0.17, p-value=0.01, permutation-test, **Supplementary Table S9**), linking neurophysiology and brain structure. This link was significant across all electrodes, with the strongest signal emerging from the F8 electrode (**Fig. 3d, Supplementary Table S9**), capturing right dorsolateral prefrontal (rDLPFC) brain activity, strongly involved in executive function and working memory^39^. Similarly, variation in EEG band power particularly in the theta and gamma range was predictive of changes in the *Cognition score* (r=0.2, p-value= 0.004, permutation-test, **Supplementary Table S9**), consistent with the hypothesized relevance of theta and gamma waves for higher cognitive functions (**Fig. 3d)**^37^. These observations are in line with the strong reduction in GMV within the rDLPFC (**Fig. 3e**) as one of the top-ranking regions within the ‘*schizophrenia neurocognition signature’* (**Fig. 1d**). GMV reduction in this region was strongly correlated with cognitive performance (**Fig. 3f**) as well as theta band power (**Fig. 3g**). These observations highligh the rDPLFC as one key area affected by alterations in synaptic density that contributes to cognitive impairment in SCZ.

Conversely, the prediction of SCZ-associated EEG frequency band power (Theta, Gamma 1 and Gamma 2) from matching iN imputed transcriptomic data (**Fig. 4a**) uncovered a significant association (r=0.17, 0.22, p-value=0.0006,0.005-0.01, permutation-test) with gamma 1 and gamma 2 across all individuals (**Fig. 4b, Supplementary Table S10**). Replication of these findings by applying the same trained model to empirically measured transcriptomes supported these observations reaching borderline significance for the correlation coefficient of the model for gamma 1 and gamma 2 (**Fig. 4b**, blue line) (p-value_gamma1_=0.079, p-value_gamma2_=0.08, permutation-test) and significance for the mean average error (p-value_gamma1_=0.04, p-value_gamma1_=0.001, permutation-test). Analysis of the predictive genes underlying this association revealed many key synaptic genes such as SHANK3 or NLGN2 and significant enrichment of key synaptic pathways **(Fig. 4c and 4d; Supplementary Table S10)**.

**Fig. 4:**
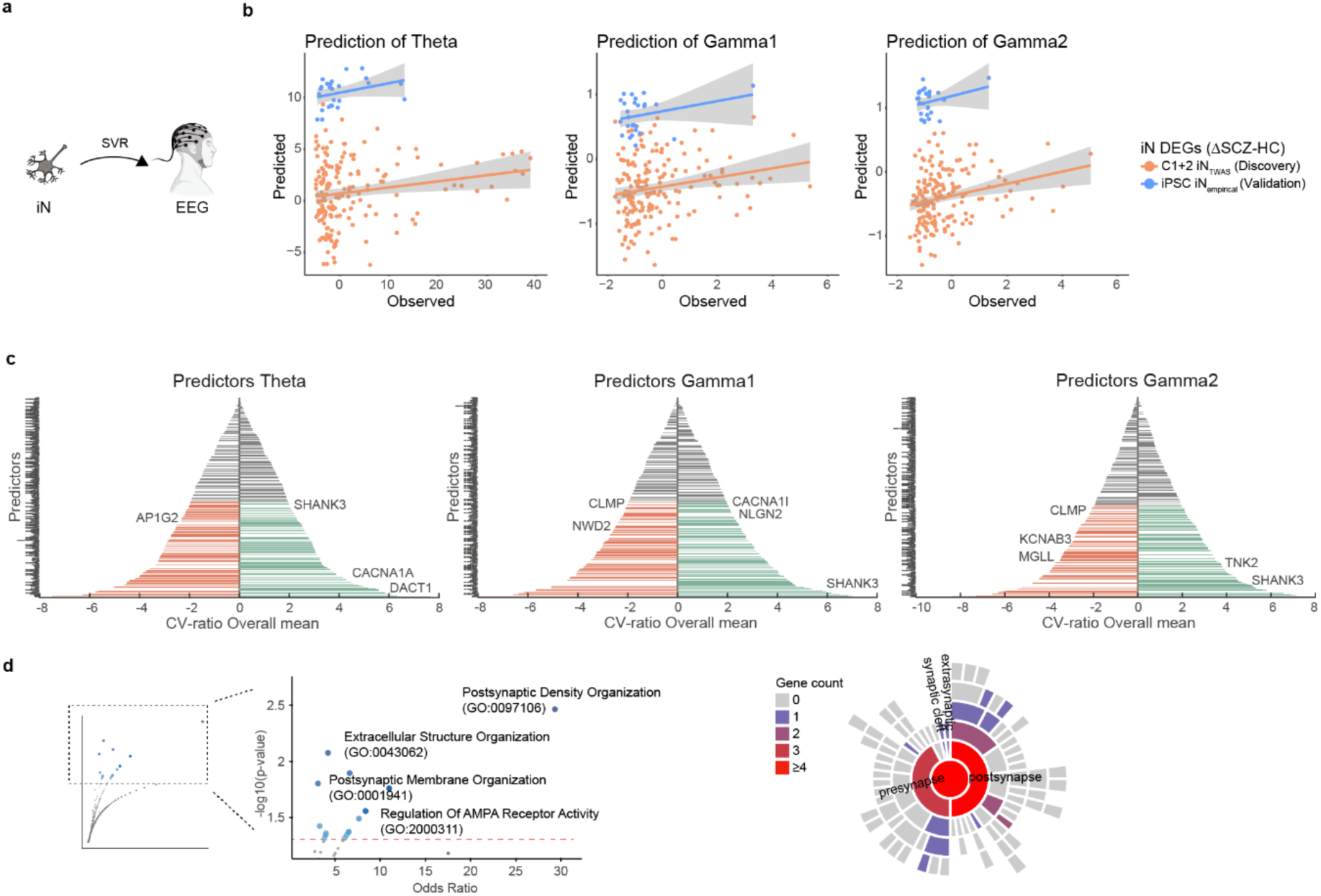
Deficits in synapse related gene expression are linked to of EEG power spectrum alterations in schizophrenia. **a,** Illustration of the prediction of the individual EEG frequency power spectra (theta, gamma1 or gamma2) using SVRs from genotype imputed gene expression profiles in iPSC derived iN of independently selected genes as features. **b,** Correlation analysis between the individual level predicted (y-axis) and observed (x-axis) absolute EEG frequency power average over all electrodes in the theta (left), gamma 1 (middle) and gamma 2 (right) band. Prediction of the band power was performed using support vector regression (SVR) from genotype imputed gene expression levels of 310 genes in iPSC derived iNs. The latter were previously identified to be differentially expressed in iPSC derived iNs and were imputed from genotype in all individuals (discovery, orange) or measured empirically in iPSC derived neurons from a subset of held out individuals (blue). The SVR model was trained using 5-fold cross validation and all predicted values are based on the held-out sample subsets. Lines indicate linear regression model for discovery cohort D (subset of cohort C1 and C2, N=178) and replication sample (C_iN_,N=26) predicting the theta (r_D_=0.18., p_D_=0.006, r_iN_=0.22., p_iN_=0.1, permutation-test), gamma 1 (r_D_=0.17., p_D_=0.01, r_iN_=0.28, p_iN_=0.08, permutation-test), and gamma 2 average power (r_D_=0.22, p_D_=0.01, r_iN_=0.28, p_iN_=0.08, permutation-test), showing the replication sample with a different offset. **c,** Feature importance plots for the SVR models in f. depicting the ranked cross-validation ratio grand means (CVR-GM, y-axis) of all gene features (y-axis) in the SVR model to predict the frequency band power for theta (left), gamma 1(middle) and gamma 2 (rights). Positive CVR-GM indicates that the feature (imputed gene) was predictive of higher band power, whereas negative CVR-GM indicates the opposite. Significant features are colored, with annotations for selected genes of interest. **d,** Left: Magnification of a volcano plot displaying gene enrichment analysis of significant CVR-OM genes that predicted absolute EEG frequency power average over all electrodes in the theta, gamma 1 and gamma 2 band using Enrichr^49,50^ based on Gene ontology (GO) Biological Process 2023^52^ with visualization based on Appyter^53^. Points on the plot represent individual gene sets with annotations for selected GO terms. The x-axis shows the odds ratio for each gene set, and the y-axis indicates the −log(p-value). Larger blue points denote significant GO terms (p-value < 0.05), while smaller gray points indicate non-significant terms. The intensity of the blue color increases with the significance of the GO term. Right: Sun burst plot illustrating locations of synaptic CVR-OM genes. Colour scheme indicates number of genes per term.

Jointly, these analyses link alterations in molecular processes contributing to synaptic function and brain structure to changes in patient level neurophysiology across different scales.

#### Cellular basis of SCZ associated changes in patient level neurophysiology

These results strongly imply altered synapse biology as one key contributor to cognitive impairment in SCZ as one phenotypic consequence. However, while our neuroimaging-based observations of structural brain alterations support a reduction in synaptic density in specific brain regions and circuits in line with previous work^6^, they lack sufficient resolution to investigate this further. Conversely, gene expression changes further support altered synapse biology as one mechanism contributing to neurophysiological changes based on EEG and ultimately cognitive impairment. However, despite these associations, there remains a substantial mechanistic gap between changes in gene expression levels, potential cellular consequences and corresponding differences in patient level intermediate and clinical phenotypes *in vivo*.

In order to narrow this gap, we hypothesized that alterations in synaptic density of patient derived *in vitro* neuronal networks constitute suitable cellular endophenotype that can be *intraindividual* translated to physiologically relevant changes in circuit micro-connectivity in individuals with SCZ.

To test this hypothesis, we leveraged biophysical neural mass models of cortical circuits and dynamic causal modeling (DCM) to predict EEG patterns based on different neuronal synaptic density parameters^18^. In contrast to previous approaches that systematically fitted synaptic density parameters *in silico* to reproduce the patient-level EEG power spectrum, we harnessed the unique features of iPSC technology. In particular, we employed iPSC derived neuronal networks to provide a direct intraindividual empirical assessment of the relative synaptic density *in vitro* across individuals with matching *in vivo* EEG data. We devised a *reverse personalized dynamic causal modeling* (rpDCM) approach, predicting the individual level EEG power spectrum using the canonical microcircuit DCM but parameterized with empirically measured synaptic density from the *in vitro* derived neuronal networks^18^(**Fig. 5a**).

**Fig. 5:**
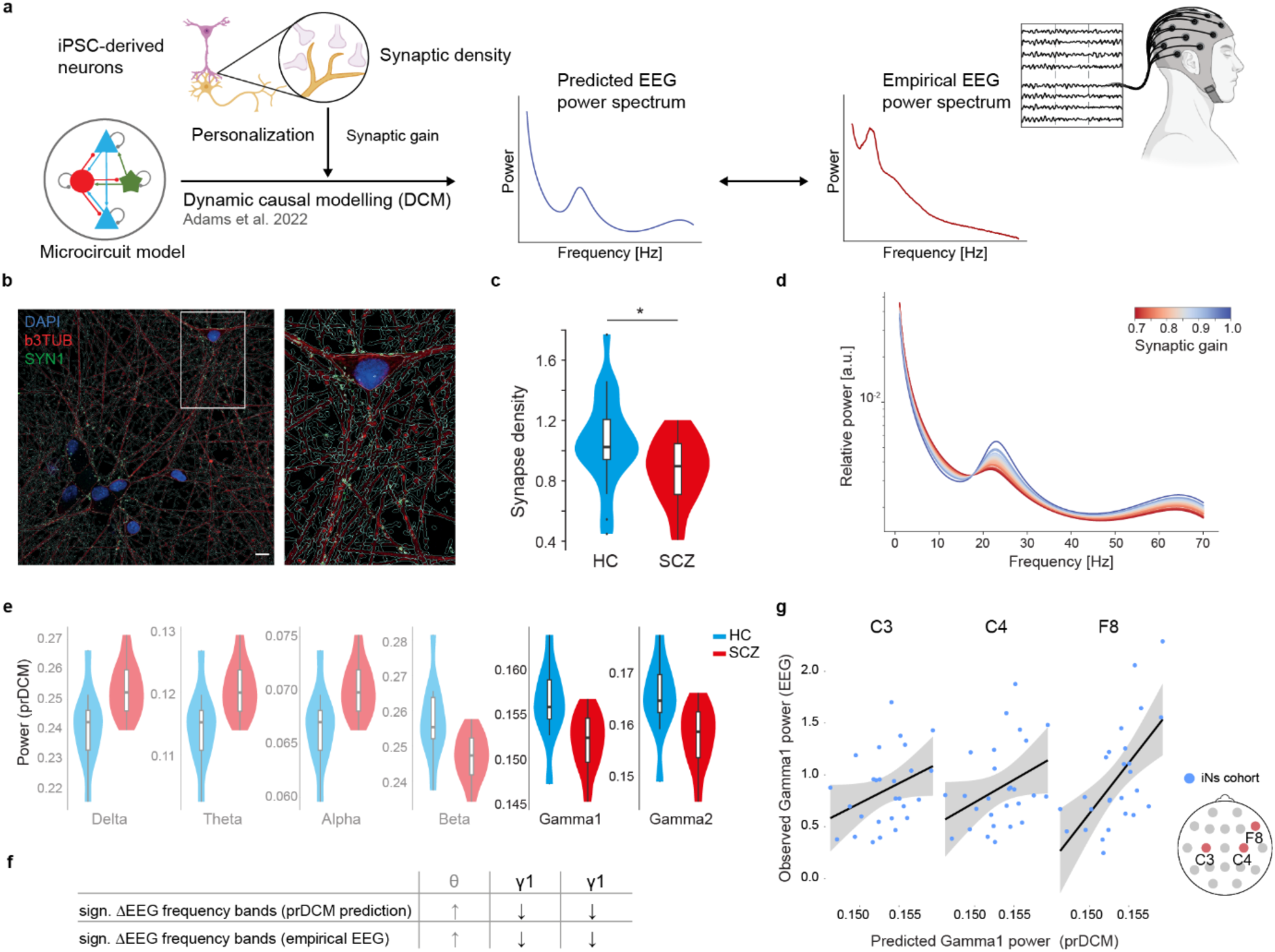
Dynamic Causal Modeling with Personalized Synaptic Density from iPSC-Derived Neurons predicts gamma power. **a,** Illustration of the iPSC-derived personalization of the dynamic causal modeling (DCM) approach, approach as developed by Adams et al. (2022) ^18^, integrating measured excitatory synaptic density from iPSC-derived neurons of patients with schizophrenia (SCZ) and healthy controls. The model uses excitatory synaptic density as a proxy for synaptic gain to generate predictive EEG power spectra for the translational EEG cohort (N=42) with available clinically measured EEG power spectrum data. **b,** Representative images from high content imaging (HCI) analysis, showcasing immunocytochemistry (ICC) signals of Synapsin 1 (SYN1) in red, β3-tubulin (β3TUBB in green), and DAPI nuclei staining in blue. The images illustrate the neuronal structures with neurite segmentation masks in iPSC-derived induced excitatory neurons (iNs). Scale bar indicates 10 µm. **c,** Violin plot illustrates the distribution of normalized SYN1 punctae density overlapping with neurites in iNs derived from healthy controls (HC) and individuals with schizophrenia (SCZ) at day 49. Density measurements were conducted using high content imaging and confocal microscopy aross iN samples from 42 individuals (N=21 HC, N=21 SCZ), encompassing a total of 107 wells/coverslips from at least two independent differentiation batches. Statistical significance of differences in punctae density between HC and SCZ is indicated by a p-value= 0.0219 from a two-tailed linear mixed model comparison. **d,** Graph illustrates the individual predicted power spectra within the translational EEG cohort (N=42) applying synaptic density as a proxy for synaptic gain within the applied DCM. **e,** Violin plots illustrate the distribution of the predicted EEG frequency band power across different frequency bands (Delta, Theta, Alpha, Beta, Gamma1, Gamma2) of the power spectrum in HC (blue, N=10) and SCZ (red, N=20). Statistical significance of differences in punctae density between HC and SCZ is indicated by a p-value from a two-tailed linear mixed model comparison. **f,** Table highlights the directional changes of the power of the different EEG frequency bands that are significant different in SCZ compared to HC in the clinically measured EEG. EEG frequency bands with significant changes in power between HC and SCZ have the same direction as personalized DCM predicts. **g,** Scatter plot showing the correlation between DCM predicted Gamma1 activity based on synaptic density from iNs of the individual and their EEG-measured Gamma1 activity of the scalp electrodes C3 (r=0.45, p=0.048, N=27), C4 (r=0.43, p=0.052, N=27) and F8 (r=0.63, p=0.0008, N=26). The trend lines and shaded areas indicate confidence intervals. Illustration on the left shows the spatial positions of the highlighted electrodes.

Therefore, we analyzed the excitatory synaptic density (**Extended Data Fig. 4a**) of highly active iN-derived neuronal networks across n=42 individuals (21SCZ/21HC) using new and previously generated^24^ measurements. This analysis revealed a significant median reduction of excitatory synapses by 12.4% in SCZ (**Fig. 5b** and **5c**; **Supplementary Table S11**). We then utilized these measurements to simulate the individual-level EEG power spectrum, setting all excitatory synaptic weights in the personalized DCM to the empirically observed relative density for each individual (**Fig. 5d**, **Extended Data Fig. 4b**). This simulated EEG power spectrum qualitatively recapitulated the groupwise differences in the power spectrum across all electrodes that were observed in the empirically measured EEG power spectrum data, including significant differences in the theta and gamma1 frequency range (**Fig. 5e** and **5f, Supplementary Table S11**). Going beyond groupwise differences, we detected nominally (p≤0.05) significant correlations between the individual level simulated and measured power in the gamma 1 range at the C3, C4 and F8 electrodes (**Fig. 5g, Supplementary Table 11**), of which the latter passed also multiple testing correction (BH corrected p-value=0.017). These results provide a biophysical link between alterations in synaptic density and changes in the gamma1 EEG power spectrum in the right DLPFC on the group as well as on the individual patient level. Moreover, these findings highlight the possibility to translate *in vitro* level cellular endophenotypes to changes in *in vivo* intermediate phenotypes.

## Discussion

In this study, we sought to narrow the translational gap and determine how disease associated changes on the molecular and cellular scale translate into alterations on neurophysiological and behavioral level. We show that a complex pattern of GMV reduction with a pronounced contribution of the right DLPFC is associated with a ‘*neurocognition signature’* in SCZ. This signature is closely linked to alterations in patient level neurophysiology based on EEG, particularly in the theta and gamma frequency range of the same patients. We then show that individual level changes in these intermediate clinical phenotypes can in part be explained by genetically driven molecular and cellular variability. Our approach enables the *intraindividual translation* of genetically driven changes in neuronal gene expression and an associated reduction in excitatory synaptic density *in vitro* into individual alterations of brain circuitry, electrophysiology and ultimately cognitive function at the patient level *in vivo* (**Fig. 1a and 1b**). Thus, these results provide a link between the molecular genetic and cellular origins of synapse reduction in SCZ to its neurobiological and behavioral consequences on the patient level.

This *generalizable strategy to dissect* diseases-relevant symptom classes such as cognitive impairments in schizophrenia might help to pave the way to identify and develop new mechanism informed treatment options.

Starting from the clinical assessment of SCZ patients and HC, this strategy identified a ‘*schizophrenia neurocognition signature’*, that links structural brain changes to a multivariate clinical phenotype. These findings imply that specific structural brain alterations that likely reflect a reduction in synaptic density contribute to cognitive deficits in SCZ.

Subsequently, we show that this signature can in part be explained by genetically driven changes in neuronal gene expression in personalized *in vitro* models from the same patients. These observations indicate a genetic contribution to these behavioral and structural brain changes. Here, iPSC derived neurons serve as a pivotal, integrating technology to reveal the molecular and cellular consequences of the underlying complex genetic risk factor profiles in each individual. Despite this unique capacity of the iPSC technology, their generation and differentiation remain a time-intensive, costly process and a bottleneck of molecular dissection^40^. By combining a large cohort of iPSC derived neuronal transcriptome profiles^24^ with machine learning^33^, we were able to mitigate this challenge by imputing the genetically determined component of gene expression levels in iNs for the vast majority of all study participants. Using this approach, we uncover a core set of dysregulated genes in iNs that are predictive of both structural and phenotypic changes that are highly enriched for synaptic pathways. Thus, these findings provide a direct link between genetically driven changes in synapse related gene expression, reduction in synaptic density *in vitro*, and alterations in brain structure and cognitive performance on the individual patient level.

To provide a neurobiological basis connecting altered brain structure and cognitive function, we show that synaptic deficits translate into altered neurophysiology. Analysis of EEG recordings indicated a strong link between alterations in the theta (4-8 Hz) and gamma (30-70 Hz) power spectrum, and structural brain alteration in the right DLPFC, fronto-parietal and fronto-temporal connection that was evident on an intraindividual basis. These regions are highly relevant for cognitive functioning and frequently affected in SCZ^41–43^. Particularly increased theta and reduced gamma power at electrode F8 (corresponding to the right DLPFC) showed a strong correlation with the severity of cognitive impairment. This finding aligns with the established role of lower gamma^38^ and higher theta oscillations^44^ and the DLPFC in cognitive processes. Importantly, these EEG alterations were also significantly correlated with the genetically driven changes in synaptic gene expression observed in the iPSC-derived neurons. These result align with the idea that gamma oscillations arise from the interplay of excitatory and inhibitory neurons, and that a reduction in excitatory synaptic function progressively contributes to localized cortical disinhibition, disrupting local E-I balance^5^. Thus, these findings provide compelling evidence that reduced synaptic density and neurotransmission contribute directly to the altered neurophysiological activity and, consequently, contribute to cognitive deficits.

To mechanistically support this link further, we devised the concept of *reverse personalized dynamic causal modelling* to biophysically model the individual EEG power spectrum based on the experimentally determined glutamatergic synapse density in iNs from patients with measured EEG data. Strikingly, this analysis reproduced all empirically observed group level differences and identified a reduction in the excitatory synaptic density in the right DLPFC region as a plausible cause of alterations in the gamma wave spectrum in SCZ. Importantly, this interpretation is further supported by the concomitant GMV reduction in the right DLPFC.

While these findings provide insights into molecular origins, neurobiological mechanisms and behavioral consequences of changes in synapse biology, many open questions remain. These include foremost the role of (fast spiking) inhibitory neurons and GABAergic synapses as key contributor to gamma oscillations and previously prominently implied in SCZ.

Similarly, the origin and role of altered excitatory and inhibitory neurotransmission remain unclear in light of previous reports on e.g. region specific increase in glutamate and decrease in GABA concentrations in brain of SCZ patients. Finally, including the neurophysiological impact of different environmental variables explicitly in this multi-scale modeling approach will be crucial to obtain a holistic understanding of disease etiology.

Addressing these questions requires further extensions of both clinical and molecular phenotyping as well as more advanced cellular model systems and computational circuit models. For the latter, our investigation was limited to changes in excitation and limited power to explain changes in the theta band. Thus, as a next step, inhibitory neurons and GABAergic signaling need to be integrated into both cell culture and circuit models, including chemical neurotransmission as a separate parameter. Moreover, the extension of translational phenotype batteries that can be assessed both *in vivo* and *in vitro* such as connectivity measurements (e.g. task and resting state-fMRI and multi-electrode arrays) or measurement of neurotransmitter concentrations (e.g. magnetic resonance spectroscopy, metabolomics) from the same individuals will be an essential prerequisite to further advance this approach. Lastly, the establishment of substantially larger translational deep phenotype cohorts with matching iPSCs available (>200 individuals) will instrumental to obtain sufficient detection power and mitigate the high level of clinical heterogeneity in SCZ.

In summary, our findings support an integrated model where genetic predisposition, acting through altered expression of synaptic genes, leads to reduced excitatory synaptic density in key cortical and subcortical regions. These cellular-level changes, potentially amplified by environmental factors, contribute to widespread structural brain alterations, likely reflecting reduction in synaptic density. These structural changes, in turn, manifest as disrupted neurophysiological activity, particularly reduced gamma band power, contributing to cognitive impairment in SCZ.

On the methodological level, this study establishes a generally applicable strategy to bridge intra-individually the gap between *in vitro* cellular endophenotypes and *in vivo* clinical intermediate phenotypes, highlighting the potential of this approach for translational research in mental health.

The combination of clinical deep phenotyping with iPSC technology, enabled us to demonstrate the potential of this *translational deep phenotyping* concept in mental health. By linking alterations in intermediate phenotypes across diverse scales, we have established a causal chain across different layers of biological complexity, ranging from genes to behavior. This intraindividual multi-layered approach underscores the power of integrating iPSC technology, machine learning and biophysical personalized dynamic causal modelling in advancing our understanding of the molecular and cellular underpinnings of clinically relevant disease phenotypes such as cognitive impairments in SCZ. By identifying potentially targetable molecular and cellular mechanisms, this approach might enable the establishment of empirically and genotype informed personal multi-layer treatment response models. These models might pave the way for target identification, and subsequent drug development that could ameliorate cognitive impairments in schizophrenia.

## Data availability

All data is available in the supplementary data or upon reasonable request to the authors.

## Code availability

All relevant code is available through the respective repository under https://github.com/zillerlab.

## Acknowledgments

This work was supported by BMBF, eMed grant numbers 01ZX1504, 01ZX1706A (MJZ), Else-Kroener-Fresenius Stiftung grant A54 (MJZ), DFG Grants GZ: ZI 1614/5-1,ZI 1614/7-1 (MJZ). The procurement of the MRI scanner was supported by the Deutsche Forschungsgemeinschaft (DFG, German Research Foundation) grant for major research (DFG, INST 86/1739-1 FUGG).

EB received funding from the Pesl-Alzheimer-Stiftung (2024-2025). DP and FJR were supported by the Else Kröner-Fresenius Foundation (Research College “Translational Psychiatry”) for their Residency/Ph.D. track at the International Max Planck Research School for Translational Psychiatry (IMPRS-TP), Munich, Germany. FJR and ECS were supported by the Munich Clinician Scientist Program (MCSP) of the Faculty of Medicine, LMU Munich, Munich, Germany (FöFoLe 009/2019 and Advanced Track 01/2021, respectively). FJR received funding from the Lisa Oehler-Stiftung (2022– 2024), the Pesl-Alzheimer-Stiftung (2024-2025). VY was supported by the Residency/PhD track of the International Max Planck Research School for Translational Psychiatry (IMPRS-TP) and was supported by the Faculty of Medicine at LMU Munich (FöFoLe Reg.-Nr. 1226/2024). JM was supported by the Faculty of Medicine at LMU Munich (FöFoLe Reg.-Nr. 1167). The study was supported by the EU HORIZON-INFRA-2024-TECH-01-04 project DTRIP4H 101188432 to PF, AS and FR. PF, AS, GH and VY received funding from the BMBF within the Era-Net Neuron project GDNF_UpReg (FKZ 01EW2206). The study was endorsed by the Federal Ministry of Education and Research (Bundesministerium für Bildung und Forschung [BMBF]) within the initial phase of the German Center for Mental Health (DZPG) (grant: 01EE2303C to AH, and 01EE2303A, 01EE2303F to PF).The study was supported by the Supplement to BMBF funding for the German Centre for Mental Health (DZPG) by the Bavarian State Ministry for Science and the Arts with the Grant for the research project ‘Improving Infrastructures for DZPG and NAKO Cohorts” to PF, DK and BK. TS received funding through the Else Kröner Medical Scientist Kolleg „ClinbrAIn: Artificial Intelligence for Clinical Brain Research” and is supported by the International Max Planck Research School for Intelligent Systems (IMPRS-IS)._AL is a member of the Machine Learning Cluster of Excellence, EXC number 2064/1 - project number 39072764.

We thank all patients for their participation making this study possible. We thank Rick Adams for sharing the DCM code and helpful discussions.

## Author contributions

Conceptualization: FJR, DP, DK, MZ; Data curation: FJR, DK, DP, LEF, LK, GH, BK, EB, LR, VY, JM, IP; Formal analysis: CV, DP, DK, SP, MZ; Wet lab experiments: VA, AT, MG; Funding acquisition: PF, NK, DK, EW, AS, MJR; Investigation: DP, DK, MZ, CV, GH, FJR, BK, EB, TJS, AL; Project administration: FJR, DK, LK. LEF; Resources: CDP, EW, SA, LR, PF, NK, MJR; Supervision: FJR, DK, MZ; Writing – original draft: FJR, MZ; Writing – review & editing: All authors

## CDP Working Group

Stephanie Behrens, Emanuel Boudriot, Man-Hsin Chang, Valéria de Almeida, Sylvia de Jonge, Fanny Dengl, Peter Falkai, Laura E. Fischer, Nadja Gabellini, Vanessa Gabriel, Sabrina Galinski, Thomas Geyer, Katharina Hanken, Alkomiet Hasan, Genc Hasanaj, Alexandra Hisch, Georgios Ioannou, Iris Jäger, Marcel Kallweit, Temmuz Karali, Susanne Karch, Berkhan Karslı, Daniel Keeser, Christoph Kern, Nicole L. Klimas, Maxim Korman, Nikolaos Koutsouleris, Lenka Krcmar, Verena Meisinger, Julian Melcher, Matin Mortazavi, Joanna Moussiopoulou, Karin Neumeier, Frank Padberg, Boris Papazov, Irina Papazova, Sergi Papiol, Pauline Pingen, Oliver Pogarell, Siegfried G. Priglinger, Florian J. Raabe, Lukas Roell, Moritz J. Rossner, Philipp Sämann, Andrea Schmitt, Susanne Schmölz, Eva C. Schulte, Enrico Schulz, Benedikt Schworm, Elias Wagner, Sven Wichert, Vladislav Yakimov, Peter Zill

## Conflict of Interest

The authors declare that there are no conflicts of interest in relation to the subject of this study. General declaration of potential conflict of interests: SG is part-time employees by and shareholders of Systasy Bioscience GmbH, Munich, Germany. MJR is shareholder and consultant of Systasy Bioscence GmbH. AH received speaker fees from AbbVie, Advanz, Janssen, Otsuka, Lundbeck, Rovi, and Recordati and was a member of the advisory boards of these companies and Boehringer Ingelheim. BS and MZ received speaker fees from Novartis Pharma GmbH. EW was a member of the advisory boards of Boehringer Ingelheim and Recordati. OP received speaker fees from Lundbeck, Otsuka, Takeda, and Janssen and was a member of the advisory boards of Lundbeck and Janssen. PF received speaker fees from Boehringer Ingelheim, Janssen, Otsuka, Lundbeck, Recordati, and Richter and was a member of the advisory boards of these companies and Rovi.

All other authors report no potential conflicts of interest.

---

**Extended Data Fig. 1:**
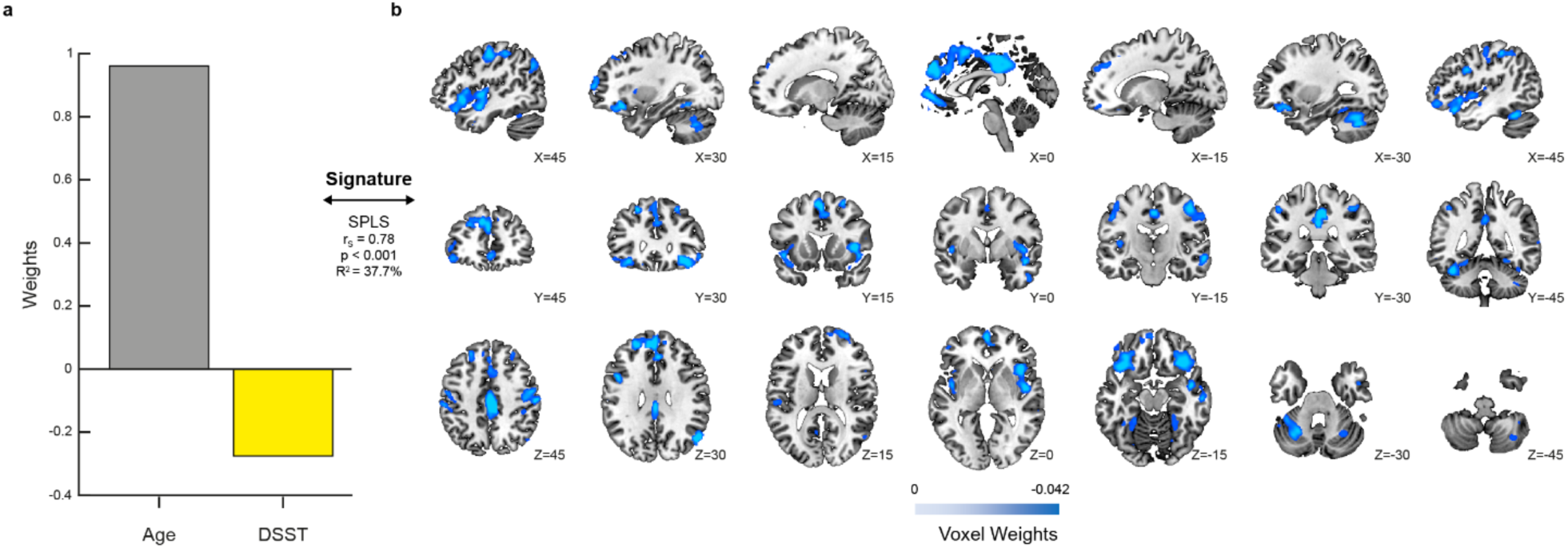
Aging signature revealed by SPLS analysis. **a,** The barplot visualizes the phenotypic pattern of the ‘*aging signature*’ contained in Latent Variable 1 (LV1), derived by the multivariate sparse partial least squares (SPLS) algorithm (N = 131, r = 0.78, p < 0.001, R^2^ = 37.7%). **b,** Illustration of the brain pattern of the ‘*aging signature*’ contained in Latent Variable 1 (LV1), derived by SPLS algorithm (N = 131). Positive weighting of voxels is indicated in the red and negative weighting in the blue color scale. The brain patterns were visualized in the MNI152 standard space using the open-source 3D rendering software Connectome Workbench v1.4.2. (https://humanconnectome.org/software/connectome-workbench).

**Extended Data Fig. 2:**
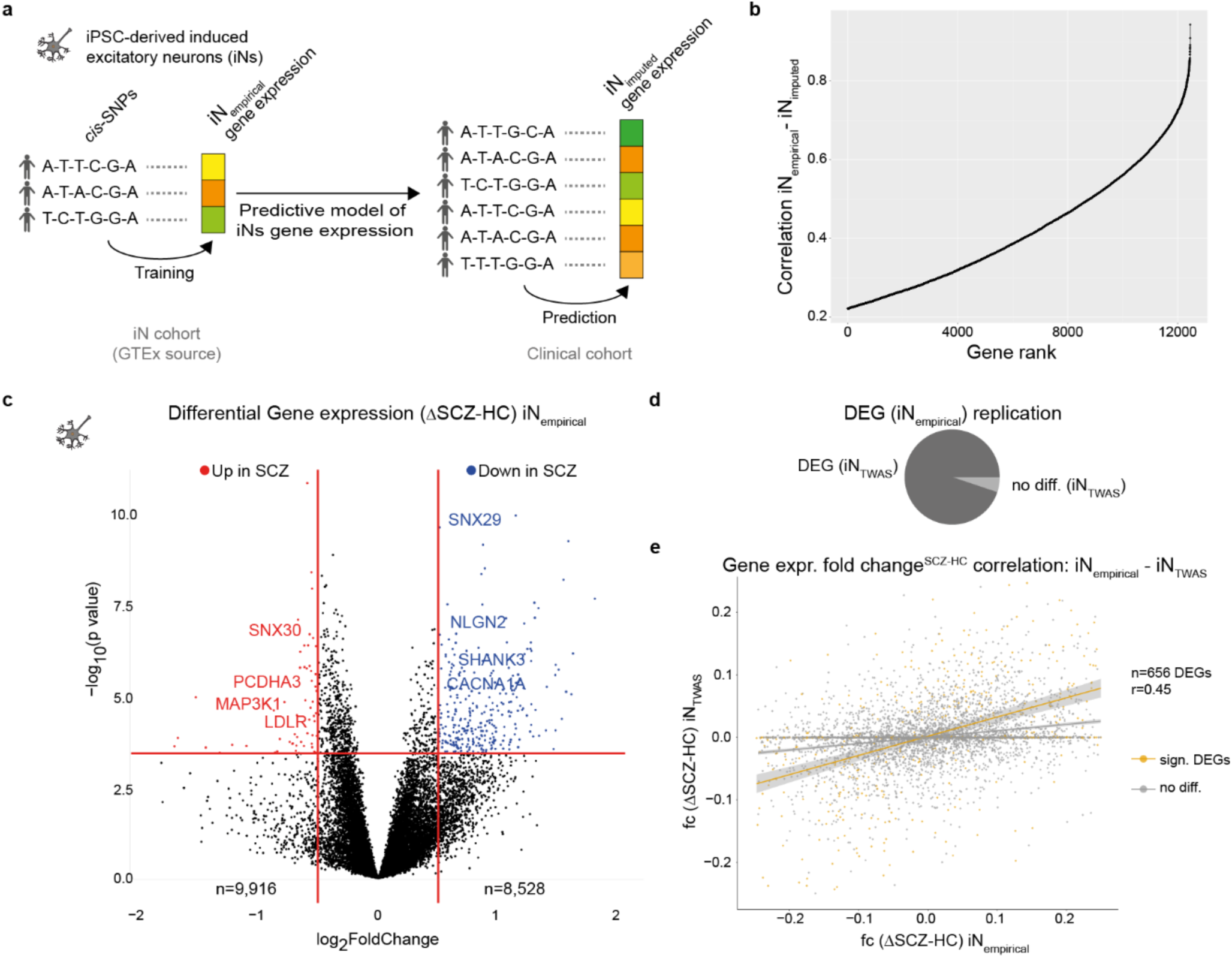
Imputed gene expression in iPSC-derived excitatory neurons and analysis of empirical and imputed differential gene expression. **a,** Scheme illustrates the development of a predictive model to impute gene expression from genetic variants in iPSC-derived induced excitatory neurons (iNs). The model is trained using a dataset from an iPSC cohort ^24^ and subsequently applied to predict gene expression in the clinical cohort based on their genotypes. **b,** Rank curve illustrates the distribution of correlation coefficients between imputed and empirically measured gene expression across the transcriptome of iNs from n=79 donors. **c,** Volcano plot showing the log2 fold-change (x-axis) and significance (-log10 p-value, y-axis) of differentially expressed genes (DEGs) between SCZ and HC iNs at day 49. Positive fold-changes indicate lower expression in SCZ. Red/blue dots indicate significance (p(FDR)<=0.01) and minimal fold-change (|fc|>=0.4) cutoffs to define DEGs. Text highlights selected DEGs. **d,** Pie charts illustrates overlap (dark grey) of significant DEGs based on transcriptome-wide association study (TWAS) analysis with the empirical DEGs (from c). **e,** Correlation of gene expression fold changes (fc) between SCZ and HC in empirical and imputed gene expression. Line in orange represents the Pearson correlation coefficient r (r=0.4468781, p-value: < 2.2e-16, n=656) for the fold change of significant differentially expressed genes (DEGs), while the dark grey line indicates the correlation coefficient for non-significant DEGs (r=0.2501, p-value: < 2.2e-16, n=4870). Each line is accompanied by a confidence interval, shaded in light grey.

**Extended Data Fig. 3:**
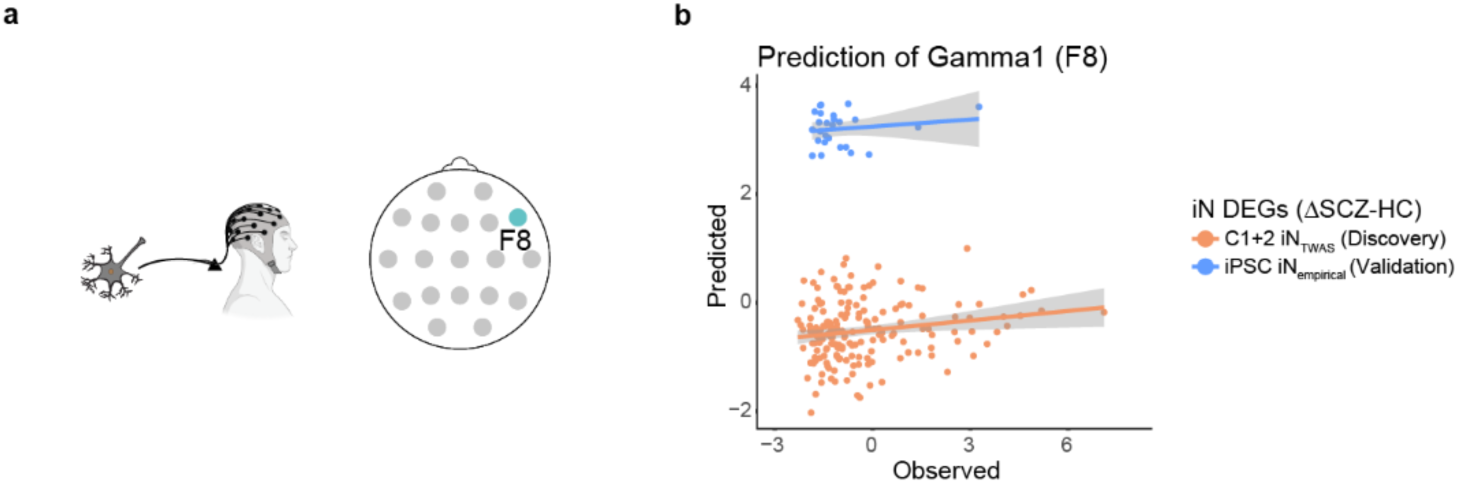
EEG power spectrum alterations of Gamma 1 frequency band relate to the individual molecular signature. **a,** Illustration of the prediction of the individual EEG frequency power spectra of Gamma 1 of F8 electrode using SVRs from genotype imputed gene expression profiles in iPSC derived iN of independently selected genes as features. **b,** Correlation analysis between the individual level predicted (y-axis) and observed (x-axis) absolute EEG frequency power average over all electrodes in the theta (left), gamma 1 (middle) and gamma 2 (right) band. Prediction of the band power was performed using support vector regression (SVR) from genotype imputed gene expression levels of 310 genes in iPSC derived iNs. The latter were previously identified to be differentially expressed in iPSC derived iNs and were imputed from genotype in all individuals (discovery, orange) or measured empirically in iPSC derived neurons from a subset of held out individuals (blue). The SVR model was trained using 5-fold cross validation and all predicted values are based on the held-out sample subsets. Lines indicate linear regression model for discovery cohort D (subset of cohort C1 and C2, N=178) and replication sample (C_iN_,N=26) predicting the gamma 1 (r_D_=0.17., p_D_=0.01, r_iN_=0.28, p_iN_=0.08, permutation-test), average power, showing the replication sample with a different offset.

**Extended Data Fig. 4:**
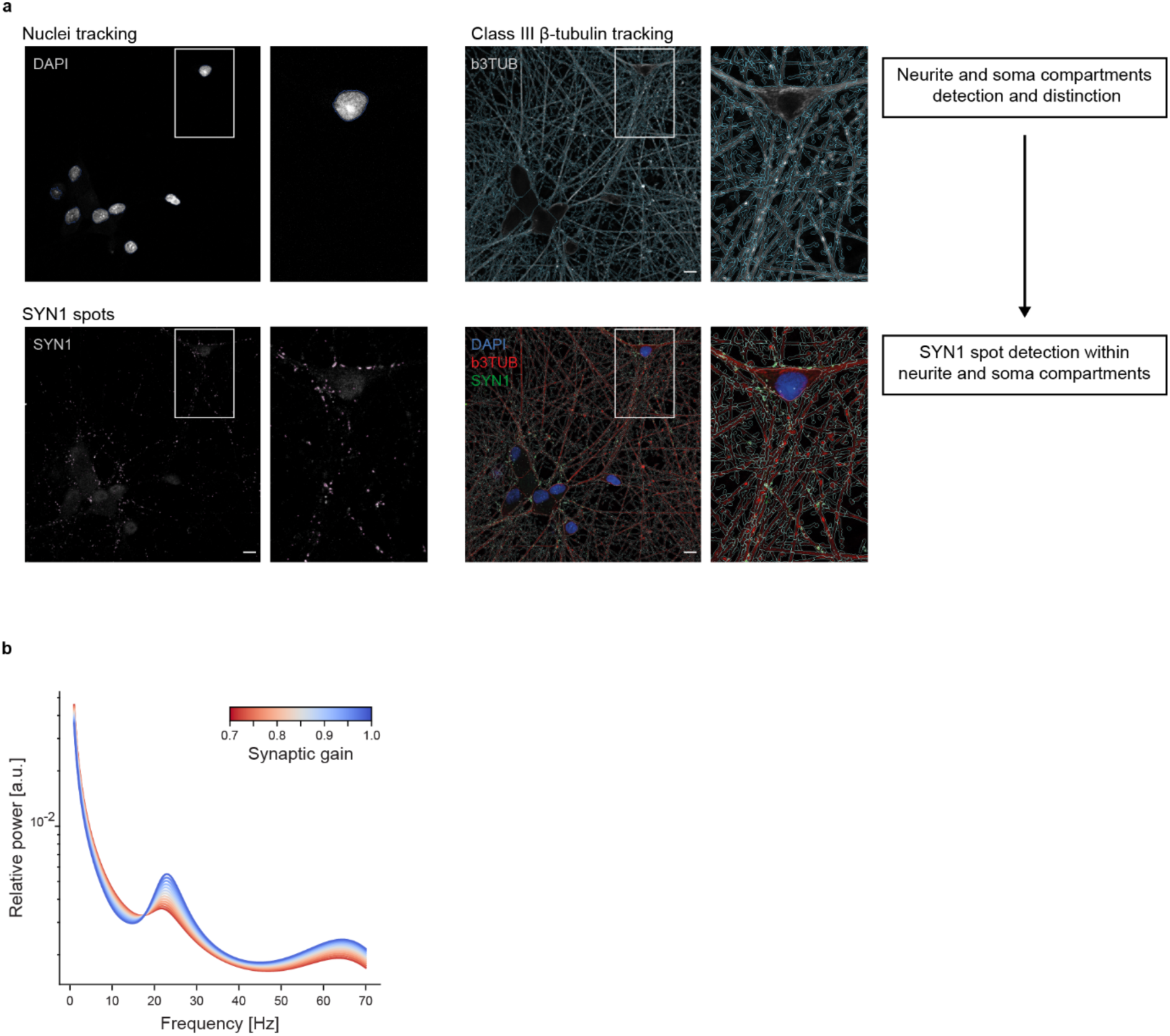
High-Resolution Imaging to determine synaptic density and Synaptic Gain Modulation in Dynamic Causal Model. **a,** Representative examples of the applied high-resolution confocal-microscopy images analysis pipeline with the original florescence channels and the respective mask. White rectangles indicate position of the respective high magnification field of view. Scale bar indicates 10 µm. **b,** Graph illustrates the application of the Dynamic Causal Model (DCM) by Adams et al. to predict the EEG power spectrum with synaptic gain changes varied in increments of 0.05. The plot shows the effect of changing synaptic gain on the relative power across the EEG power spectrum, indicated by the blue and red lines which represent different levels of gain adjustment.

## Online Methods

### Participant Recruitment and definition of analysis groups

Participants of Cohort 1 (C1) were part of the *‘Multimodal Imaging in Chronic Schizophrenia Study’* (MIMICSS) that was performed at the Department of Psychiatry, University Hospital, LMU Munich, Germany (see supplements from^1^, and^2^). This study was approved by the local ethics committee of Faculty of Medicine, LMU Munich (project number: 17–13). The patients who provided written informed consent were diagnosed by two independent, experienced psychiatrists using the criteria of the International Statistical Classification of Diseases and Related Health Problems, 10th revision (ICD-10). Beside first-degree unaffected relatives (URs), also unaffected healthy controls with no current or past mental illness according to the Mini International Neuropsychiatric Interview, M.I.N.I.^3^ were recruited.

Cohort 2 (C2) originates from the Clinical Deep Phenotyping study, study protocol was described in details elsewhere ^1^, which is a multimodal add-on study of the Munich Mental Health Biobank (ethics project number 18-716)^4^. The study was registered at the German Clinical Trials Register (DRKS, ID: DRKS00024177) and received approval from the local ethics committee of the Faculty of Medicine, LMU Munich (project numbers 20-0528 and 22-0035). It encompasses patients as well as HCs who had no lifetime psychiatric disorders as determined by the Mini International Neuropsychiatric Interview ^3^. For this study, we included only patients with schizophrenia spectrum disorders (SSD), mainly with SCZ, URs and HCs from C1 and C2 were included. Psychotic symptom severity was determined with the Positive and Negative Syndrome Scale (PANSS)^5^. Cognitive performance in C1 was determined with by trained study staff with several task covering diverse domains of cognitive performance including the Verbal Learning Memory Test ^6^ and the Trail Making Test ^7^. For C2, the Brief Assessment of Cognition in Schizophrenia (BACS)^8^ was performed. Current antipsychotic treatment levels were was converted to chlorpromazine equivalent doses (CPZeq)^9^. Further study details from C1 and C2 along are available in the **Supplementary Table 1**. Detailed list of all participants included in each of the different analyses can be found in the corresponding Supplementary Tables.

### Genotyping, quality control, imputation, and calculation of polygenic risk scores

Individuals were genotyped using Illumina’s Global Screening Array (Life & Brain GmbH). Illumina GenomeStudio v2.0.4 (llumina Inc., San Diego, CA, USA) was used to process global screening array genotyping data and to obtain genotype calls for the study participants. Quality control (QC) was performed using PLINK v1.9/v2^10^. Single-nucleotide polymorphisms (SNPs) were excluded if they had a missing call rate greater than 2%, had a Minor Allele Frequency (MAF) less than 0.5%, or deviated from Hardy-Weinberg equilibrium with p < 0.0001. Individuals were excluded if they had a missing call rate greater than 2%, were duplicated samples according to the pairwise identity by state method, had a large deviation in their heterozygosity value (± 3.90 standard deviations), or had non-European ancestry according to a Multidimensional Scaling (MDS) analysis. MDS analysis was carried out with PLINK v1.9 to obtain a representation of genetic ancestry in our study, extracting the first 10 ancestry components. Palindromic SNPs and SNPs with a large MAF deviation (greater than 10%) with respect to 1000 Genomes European reference populations were also removed. Imputation was performed using the Haplotype Reference Consortium panel^11^ in the Michigan Imputation Server^12^. A post-imputation QC was carried out to exclude SNPs that had an imputation quality score of R2 less than 0.3; or had a MAF less than 1%.

After genotype imputation, genotype dosage data were used to calculate schizophrenia polygenic risk scores (SCZ-PRS) for the N=277 individuals included in the analyzed cohort, based on the results of the Psychiatric Genomics Consortium wave 3 SCZ genome-wide association study^13^. Posterior single nucleotide polymorphism effect sizes were inferred under continuous shrinkage priors using PRS-CS^14^. The global shrinkage parameter (φ) was estimated using a fully Bayesian approach. The SCZ-PRS were eventually then calculated with PLINK v1.9^10^ as the sum of risk alleles across SNPs carried by each individual multiplied by the inferred effect sizes.

### Magnetic resonance imaging (MRI)

MRI scans in C1 were conducted with a 3.0 T MR scanner (Magnetom Skyra, Siemens Healthcare, Erlangen, Germany) equipped with a standard 20-channel phased-array head coil. MRI recordings in C2 were performed on a 3T Siemens MAGNETOM Prisma scanner (Siemens Healthineers AG) with a 32-channel head coil. T1-weighted scans were acquired by using a magnetization-prepared rapid acquisition gradient-echo sequence with an isotropic voxel size of 0.8 mm^3^, 208 slices, a repetition time of 2500 ms, an echo time of 2.22 ms, a flip angle of 8, and a field of view of 256 mm^2^.

### Electroencephalography (EEG)

Resting state EEG data were collected using 32 scalp electrodes and recorded with a BrainAmp amplifier (Brain Products, Martinsried, Germany). The setup had a sampling rate of 1000 Hz, using the Cz electrode as the reference. Electrode skin impedance was maintained below 5 kΩ. The electrodes were placed according to the International 10/20 system. The EEG recording session lasted a total of 10 minutes, divided into 5 minutes with eyes closed (eyes were opened for 3 seconds every 120 seconds to mitigate excessive alpha power), followed by 5 minutes with eyes open. Throughout the session, participants were asked to stay calm and relaxed. An automated ICA preprocessing pipeline, adapted to Adams et al. ^15^ was utilized for preprocessing resting-state EEG with eyes closed condition, implemented in MATLAB (The Mathworks Inc.) using EEGLAB v2022.0 ^16^(https://sccn.ucsd.edu/eeglab/). First, the data was re-referenced to the mastoid bones (TP9 and TP10) and then downsampled to 256 Hz. A data filtering was applied, setting the frequency range between 1 to 70 Hz. rsEEG data were postprocessed in MNE Python^17^. The canonical power spectrum density was calculated using the Welch Method, with 6-second windows and 3-second overlaps. (time_frequency.psd_array_welch). YASA library was used to extract absolute and relative power ^18^ of the different frequency bands: δ (1-4 Hz), θ (4-8 Hz), α (8-12 Hz), β (12-30 Hz), γ1 (30-50 Hz), γ2 (50-70 Hz).

### iPSC culturing, neuronal differentiation

iNeuron (iN) differentiation was carried out as described previously^19^. Six-well plates were coated with Geltrex (Geltrex in DMEM/F12) on Day 0 and left at 37°C for at least an hour. After incubation with Accutase for five minutes at 37°C, iPSCs were diluted 1:1 in DMEM/F12 supplemented with 1% FBS, centrifuged for five minutes at 300g, and then resuspended in StemMACS iPS Brew XF supplemented with Polybrene (6µg/ml) and 1x RevitaCell. Cells were counted and infected with pTet-O_Ngn2-puro and FUW-M2rtTA, and firstly they were incubated for 10 minutes at 37°C at a density of 1,000,000 cells/ml, and then seeded (25,000 cells/cm²) on the Matrigel-coated plates.

The following day (Day 1), media was changed with KSR media (KO DMEM, 15% Knockout Serum Replacement, 1x NEAA, 1x Glutamax, and 50 µM beta-mercaptoethanol).

To initiate neuronal patterning and directed differentiation, the media was supplemented with 10 µM SB431542, 2 µM XAV939, 0.1 µM LDN-193189, and 2 µg/ml doxycycline.

On day 2, media was replaced with a 1:1 mixture of KSR and N2 media (DMEM/F-12, 1x Glutamax, 3mg/ml glucose and 1x N-2), supplemented with 5 µM SB431542, 1 µM XAV939, 0.05 µM LDN-193189, 2 µg/ml doxycycline and 10 µg/ml puromycin, for selection purposes.

At day 3, media was changed with N2 media supplemented with 2 µg/ml doxycycline and 10µg/ml puromycin.

At day 4, neural progenitors were dissociated with Accutase for 5 minutes at 37°C. The cell suspension was diluted 1:1 with DMEM-F12 supplemented with 1%FBS and fully dissociated to a single-cell solution.

Cells were seeded at a density of 60.000 cells/cm² in NBM media (Neurobasal medium, 1x Glutamax, 1x NEAA, 1x B27 without vitamin A, 3 mg/ml glucose, 2% fetal bovine serum (FBS), freshly supplemented with 10 ng/ml BDNF, 10 ng/ml CNTF, 10 ng/ml GDNF, and 2 µg/ml doxycycline) on plates coated overnight with Poly-L-Ornithine (15 µg/ml) and then washed and coated with laminin (1 µg/ml) and Fibronectin (2 µg/ml) for at least 4h. At day 7, 5000/cm2 primary murine astrocytes were seeded on the iN culture. To prevent glial cell division and enrich post-mitotic neurons, NBM media was supplemented with 4 µM AraC on days 10 and 14.

With a density of 5000 mouse astrocytes/cm² on the iNs, murine astrocytes were introduced by full media change on Day 7. To prevent glial cell division, 4 µM AraC was introduced to NBM medium on days 10 and 14. Afterwards, half media was removed changed every 3-4 days. From day 24 onwards, the cells were maintained in BrainPhys media (BrainPhys Neuronal Medium, 1x Glutamax, 1x NEAA, 1xB27 without vitamin A, 3 mg/ml glucose, 3% fetal calf serum (FCS), freshly supplemented with 10 ng/ml BDNF, 10 ng/ml CNTF, 10 ng/ml GDNF and 1µg/ml laminin) to promote neuronal maturation. Cells were collected at day 49 for endophenotyping.

### Imaging and synapse density analysis

A subset of synapse density measurements using the same differentiation protocol was taken from our previous publication^20^. This dataset was extended for N=31 additional donors or donor replication using the following strategy: Analysis of Syn1 density was performed by confocal imaging using Leica DMi8 with Leica Application Suite X Software of iPSC derived neurons cultured on coverslips. Images were taken at 40X magnification and a resolution of 2048 dpi and exported as a TIFF file.

For each Field of View (FOV) ImageJ was used to identify the cell bodies on the TUBB3 channel, using the freehand selection tool. The TIFF files for each channel and the soma file were imported in CellProfiler 4.2.1 and analysis was carried out in a batch-specific manner. Nuclei were detected in the DAPI file by Adaptive Otsu three classes thresholding method for intensities (>0.006) and size (40-90 pixels). Afterwards, nuclei were filtered by texture and subsequently used together with the TUBB3 file to identify cell bodies using the Distance – B method (maximum distance from detected nuclei to TUBB3 signal: 40 pixels). The final cell body mask was generated by combining the manual and automatic detected soma.

A partial neural network was identified using the Adaptative Minimum Cross-Entropy thresholding method (for intensities >0.01) in the TUBB3 mask. The cell body mask was subtracted from the partial neuronal network mask to generate the neurite mask.

Syn1 channel was adjusted with the “Speckle enhancement” function and subsequently used for identification of Syn1 speckles by Adaptive Sauvola method. Syn1 speckles were counted and quantified by occupied area (Syn1 area) in cell bodies and neurites, and normalized over the total cell body and neurite area. Next, Syn1 density measurements based on speckle area across were averaged across at least 8 FOV per well/each coverslip. Finally, Syn1 density measurements were normalized by batch, calculating a batch specific normalization factor. All results were merged with the previously published synaptic density measurements giving rise to a total of 107 distinct wells/coverslips across 4 differentiation batches and 42 different iPSC donors (**Supplementary Table 11**). To test for differences in Syn1 density between cases and controls, we performed linear mixed model analysis on Tukey’s outlier test filtered synaptic density measurements with the following formula *Neurite_Syn1Area∼ 1 +platform +(1|donor)+diagnosis* and report the results in **Fig 5c**.

To perform EEG and synaptic density association and DCM analysis, Syn1 density based on Syn1 area in the neurite compartment was averaged across all wells/cover slips for each donor (**Supplementary Table 11**).

### Imputed gene expression

We used PriLer^21^ train a machine learning model for each gene, predicting gene expression levels in iPSC derived neurons from genotype only. We utilized a cohort of previously published matching genotype and RNA-Seq data from iPSC derived pyramidal neurons from n=79 distinct donors to train individual elastic-net models for n=xx genes based on all single nucleotide polymorphisms (SNPs) within 200kb of the TSS of each gene as previously described. These models predict the variance in neuronal gene expression attributable to cis-acting genetic factors. We then applied these trained models to predict the cis-component of all genes across the discovery and replication cohort. In order to also account for trans-acting genetic factors, we next established a support vector regression model to predict the gene expression levels of n=310 genes from both cis- and trans-factors. The latter genes were identified previously to be differentially expressed between SCZ and HC iPSCs derived neurons (**Supplementary** Fig. 2c). To this end, we trained a support-vector regression model using the e1071 toolbox with a linear kernel, inferring the optimal hyperparameters using 5-fold cross validation. Prediction of the 310 candidate gene expression levels for all individuals with genotype included in this study was performed from all (cis-based) imputed transcriptome features, pre-filtering the genes for association with diagnosis status below a p-value of 0.1 to reduce the feature space. These analyses were performed fully independent for the discovery or replication cohorts (**Supplementary Table 1**). The predicted expression levels of these 310 genes showed good agreement with the empirical data (**Supplementary** Fig. 2b,d) and were used as input features for subsequent prediction of brain/behavior score or EEG patterns.

### Dynamic causal modelling

For the dynamic causal model (DCM), we adapted a previously established canonical microcircuit model^15^, representing a biophysical model of interacting pyramidal, interneuron, and spiny stellate Neuronal cell populations (**Fig. 5a)**. This model simulates neuronal mass and resulting circuit activity, giving rise to a simulated EEG power spectrum. The model can be parameterized with different neuronal connectivity settings and strength within and between the various neuronal populations to assess the impact of these parameters on the EEG. We parameterized the model by the synaptic strength (synaptic gain) of all excitatory synapses in the microcircuit and simulated the resting state EEG pattern as a function of different overall synaptic strength levels. Subsequently, utilized our empirical synaptic density measurements in iNs obtained from 42 SCZ and HC donors (**Fig. 5b,c**), normalized them to the synaptic gain parameter interval of the DCM ([0.7, 1.0]) to obtain personalized DCMs for each SCZ patient and HC. Subsequently, we simulated the respective relative EEG power spectrum of each individual (**Fig. 5d,e**) and compared the absolute power for each frequency range to the power of the eyes closed, resting state EEG for each individual and electrode. Association analysis between the observed and predicted power across individuals with matching empirical EEG and personalized DCM based EEG across all electrodes identified the simulated gamma1 power for the F8 electrode to be significantly predictive of the empirical power after multiple testing correction (q-value=0.017, T-Test), while a similar pattern was observed for the C3 and C4 electrodes at nominal significance.

### Statistical analysis

Group comparisons and association analyses were performed using linear (mixed) or logistic regression models as indicated in the respective sections. Results were corrected for multiple testing using q-value as implemented in the respective R package^22^ where appropriate and described in the respective analysis.

#### Sparse partial least squares algorithm

The SPLS algorithm used in this analysis follows the original publication of Monteiro et al^2^ and has been described in a previous publication of our group^3^. Specifically, we used the open source toolbox by Popovic et al^8^. Like Partial Least Squares (PLS), SPLS requires two data matrices *X* and *Y* as inputs. In our study, *X* contains structural MRI data, while *Y* contains cognition (BACS) and additional features (age, sex, years of education, study group status, IQR). *n* is the number of samples; *p* is the number of features in *X* and *q* is the number of features in *Y*. PLS provides insights into the brain’s mechanisms by finding relationships between different measures (i.e., views) from the same participants, i.e., between neuroimaging and cognitive data, in a given population. PLS identifies a projection or latent space containing the relevant information in both views by finding pairs of weight vectors (generally called *u* and *v*) which maximize the covariance between the projections of the two views^9^:

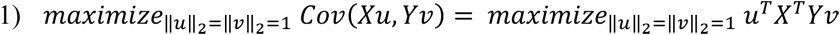

The weight vector pair is also called a latent variable (LV) as it explains one specific associative effect between the two different views. More specifically, the weight vectors place weights on each feature in the cognitive and the neuroimaging dataset, thus visualizing which features are associated with each other as well as the direction and the strength of this multivariate association. Hence, by studying this latent space, one can learn about the underlying relationship between cognitive information and brain measures_2._

In contrast to regular PLS, SPLS enforces sparsity on the weight vectors *u* and *v* through hyperparameters *c*_*u*_ and *c*_*v*_. *c*_*u*_ and *c*_*v*_ are the regularization hyperparameters that control the *l*_1_-norm constraints of *u* and *v*, respectively. The *l*_1_-norm constraints impose sparsity, which means that the lower the values of *c*_*u*_ and *c*_*v*_ are, the higher the sparsity in the respective view is ^10^. This leads to the following optimization problem:

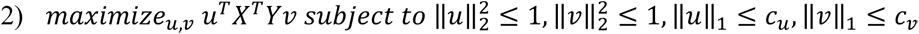

Yet, this type of constraint can only select up to *n* features if *p* > *n*. Furthermore, it will remove features which might be relevant for the model but are correlated with other features which are already included. Zou and Hastie addressed this issue by adding the *l*_2_-norm constraints^11^. For both *l*_1_-norm and *l*_2_-norm constraints to be active, the values of the hyperparameters must be between 1 and the square root of the number of features in the respective matrices. Therefore, the hyperparameter space is updated:

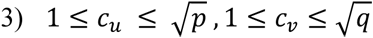

Using the hyperparameter space of equation 3) and solving the optimization problem of equation 2) according to Witten et al. ^10,12^ leads to the following SPLS algorithm steps as described in Monteiro et al.^2^:

1. Let *C* ← *X*^*T*^*Y*
2. Initialize *v* to have ‖*v*‖_2_ = 1
3. Repeat until convergence:

a. Update *u*:

i. *u* ← *Cv*
ii. 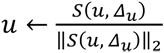, where *Δ_u_* = 0 if this results in ‖*u*‖_1_ ≤ *c_u_*, otherwise *Δ_u_* is set to be a positive constant such that ‖*u*‖_1_ = *c*_*u*_
b. Update *v*:

i. *v* ← *C*^*T*^*u*
ii. 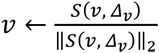, where *Δ*_*v*_ = 0 if this results in ‖*v*‖_1_ ≤ *c*_*v*_, otherwise *Δ*_*v*_ is set to be a positive constant such that ‖*v*‖_1_ = *c*_*v*_
4. If convergence is not reached after the iteration limit (default: 1000), return non-sparse weight vectors *u* and *v*

After a weight vector pair (*h*) is found by SPLS, its effect needs to be removed from the data, to look for the next possible weight vector pair (*h* + 1). This process is called matrix deflation. In this setup, projection deflation is used as it has been shown to outperform the classic Hoteling’s deflation, which is also used in Principal Component Analysis^2,13,14^. For matrices *X* and *Y*, the deflation process from iteration *h* to iteration *h* + 1 is therefore computed as follows:

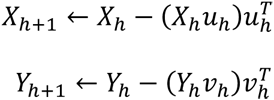

The algorithm then uses the deflated matrices and looks for the next associative effect, i.e., the next LV. This way, SPLS iteratively provides LVs consisting of sparse weight vector pairs (*u*, *v*), uncovering several layers of associative effects within the dataset.

The second step of the SPLS algorithm involves the creation of latent scores. For every LV, weight vectors *u* and *v* are projected onto the matrixes *X* and *Y*, thus generating latent scores ε and ω.

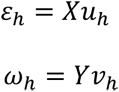

These latent scores are finite numerical values, which represent the loading of each individual on these weight vectors, e.g., how high their GM probability in certain voxels is. Therefore, every individual can be represented within each LV space with its latent pheno and brain scores. These specific scores can then be used for post-hoc analyses to investigate the meaning and relevance of these individual loadings. The models were generated and tested in a nested cross-validation (NCV) framework with 5 outer (*X*2, *Y*2) and 5 inner folds (*X*1, *Y*1). Individuals were stratified to the fold structure according to study group, so that all inner and outer folds contained an equal distribution of the three study groups (HC, UR, SCZ) to avoid training on diagnosis-related effects. Within the inner folds, a 100×100 point grid search of both hyperparameters was conducted covering the entire hyperparameter space, in which both *l*_1_- and *l*_2_-norm constraints are fulfilled: 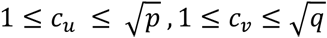 (with *p* features in matrix *X* and *q* features in matrix *Y*). Lower *c*_*u*_ and *c*_*v*_ values lead to a sparser solution, whereas higher *c*_*u*_ and *c*_*v*_ values amount to a denser solution. At the upper limit, the maximum values of hyperparameters are: 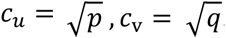. A SPLS analysis with *c*_*u*_ and *c*_*v*_ reaching these maximum values is equal to a regular PLS analysis, where every feature receives a weight and no feature is removed, i.e., no zero weights are given. Hence, our hyperparameter grid search includes the computation of one regular non-sparse PLS model (with *c*_*u*_ and *c*_*v*_ at the maximum limits) and an array of sparse PLS versions as lower *c*_*u*_ and *c*_*v*_ values are tested. Therefore, in this framework, the non-sparse regular PLS solution competes against the sparse PLS solution in the hyperparameter optimization process. The weight vector pairs were generated using the training folds in the inner loop (*X*1_*train*_, *Y*1_*train*_):

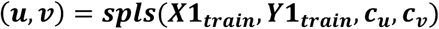

The model fit of the weight vector pair was then assessed by projecting them onto the testing folds (***X*1**_***test***_, ***Y*1**_***test***_) in the inner loop and computing Spearman’s correlation coefficient between the projections of the weight vectors ***u*** and ***v*** onto their respective data matrices ***X*1**_***test***_ and ***Y*1**_***test***_:

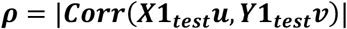

This approach delivers a simple and transparent measure of how well the weight vectors align the matrices to each other, i.e., how well they can maximize the covariance. The median correlation coefficient was computed for each hyperparameter combination in the inner loop. Afterwards, the best hyperparameter combinations (***c***_***u***-***top***_, ***c***_***v***-***top***_) with the highest median correlation coefficients (***ρ***_***top***_) were retrained on the entirety of all 5 folds of the inner loop to increase the sample size for training once more:

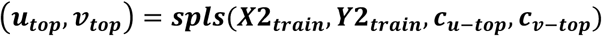

The generalizability of the weight vector pairs (**u**_**top**_, **v**_**top**_) was tested by assessing the fit of their projections onto the previously held-out fold in the outer loop and thus computing the corresponding correlation coefficients (***ρ***_***max***_).

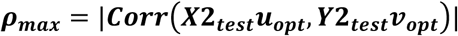

Significance testing of this weight vector pair was achieved by permutation testing against ***B*** permutations. Within the fold structure of the outer loop, ***B*** permutated datasets were created by randomly reshuffling the order of participants in one matrix (***Yb*2**) thus destroying relationship between the two matrices. The final model with the optimized hyperparameters (***c***_***u***-***opt***_, ***c***_***v***-***opt***_) was then retrained and tested in each of the ***B*** permuted datasets, thus generating weight vectors ***u***_***b***_, ***v***_***b***_:

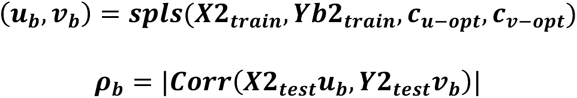

Significance testing of the LV was done by assessing how often the model based on the permuted dataset performed better or equal to the model trained on the original dataset:

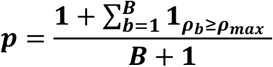

As our framework consisted of 5 outer folds, this approach led to 5 different models (i.e., 5 weight vector pairs u and v) for each LV iteration. Of these 5 different models, we selected the one model with the best performance as measured by means of permutation testing, i.e., the model that exhibited the lowest *P* value. If this optimal model passed significance testing against the FDR-corrected *P* value for multiple testing (5 models = 5 tests), the LV was deemed significant and the next LV was computed. This concept is known as the omnibus hypothesis, which was also applied in the original method paper of the SPLS algorithm.^2^ The SPLS algorithm is an iterative process, in which based on hyperparameters *c*_*u*_ and *c*_*v*_, the weight vectors *u* and v are computed in dependence of each other. First, *u* and *v* are initialized as non-sparse weight vectors based on regular singular value decomposition. Then an iterative process is set in motion, where first an enforcement of sparsity is attempted on weight vector *u* in dependence of weight vector *v*. Then sparsity is enforced on *v*, based on the previously computed weight vector u. This iterative process is repeated, where *u* and *v* are sequentially updated based on each other’s previous modification until convergence between the vectors is reached. Hence, every hyperparameter setup *c*_*u*_ and *c*_*v*_ leads to a unique process of finding converging weight vectors *u* and *v* that were generated in a dialectic manner. Thus, the multivariate information is contained in this highly specific combination of weight vectors *u* and *v*, with both vectors containing mathematical information of the other. This, in turn, makes weight vectors *u* and *v* from different models, such as in our 5×5 fold NCV, not suitable for usual merging techniques (weighted mean/mean/median merging or majority voting) as every vector *u* is dependent on the corresponding vector *v*. Therefore, we used the omnibus hypothesis to determine our final LV model out of the 5 computed within the NCV structure of each LV iteration. Using our 5×5-fold outer and inner cross-validation loops can lead to high variance in the results. After training on the inner loops and then testing on the outer loops, 5 models with 5 *P* values are obtained. A criterion is then needed to determine whether any statistically significant effects were indeed found. For this, we used the omnibus hypothesis, where a statistical test is performed *j*-times to test a null-hypothesis *H_j_.* Following the omnibus approach, the combined hypothesis H_R_ over all tests j is: “All the hypothesis H_j_ are true”. This hypothesis will be rejected if any of the *H_j_* hypothesis is rejected.^15^ In our specific case, the omnibus hypothesis states that if any of the 5 *P* values (obtained in the 5 outer folds) is statistically significant (corrected for multiple testing *j*-times), then the omnibus hypothesis will be rejected, and the detected effect will be deemed significant. Therefore, the omnibus hypothesis will be rejected if any of the 5 splits generates a *P* value below .05 (adjusted for multiple testing). Of all significant splits, the model with the lowest *P* value will be determined as the final LV model.^2^ The computation ends as soon as none of the 5 splits of the LV iteration did not pass the test for significance, which renders the entire LV not significant. Since deflating the data matrices of non-significant effects would be not justified, the analysis pipeline stops after the first non-significant LV was detected.

Full SPLS analysis was performed on the MIMICS cohort (cohort 1) as discovery sample to identify LVs. For replication of the SPLS signatures in the CDP cohort (cohort 2), propensity score matching was employed first^23,24^. Specifically, a general linear model with a binomial distribution was fitted using the matching features age, sex and study group (HC, UR, SCZ) to predict cohort assignment (MIMICSS, CDP). After that, individuals from the CDP cohort were drawn based on k-nearest neighbor computation (k=7) to retrieve a subsample of CDP individuals that most closely matched the MIMICS cohort with regards to the matching features. Following this procedure, a total of 153 individuals from the CDP cohort were drawn for SPLS model replication. Replication analysis was performed by projecting the data of the CDP sample onto the LVs retrieved in the Mimics sample and computing the correlation coefficient between the resulting latent scores. Covariate correction and sites remained the same in the CDP sample.

#### Support Vector Regression

The entire machine learning analysis was conducted on the NeuroMiner machine learning platform,^6^ release version 1.3 (https://github.com/neurominer-git/NeuroMiner_1.3).

##### Repeated nested cross-validation (RNCV)

We embedded the algorithm in a repeated nested cross-validation (CV) framework with 10 folds and 10 permutations on the inner (CV1) and outer (CV2) cycle to prevent overfitting and information leaking, while enhancing generalizability.^4^ Hyperparameter optimization was performed on the CV1 cycle, while model testing was done on the CV2 cycle.^16^ Thus, all steps involving group-level statistical procedures (e.g., scaling, pruning, covariate correction) took place in the CV1 cycle. We extended the NCV to a repeated NCV approach^4^ at both the CV2 and the CV1 level by randomly permuting the individuals within their groups and repeating the CV cycle for each of these permutations to avoid batch effects and, again, increase generalizability.

##### Preprocessing

Each feature was scaled, pruned and, if necessary, corrected for covariates within the NCV scheme.^17,18^ Only EEG data, whether used as predictors or labels, was corrected for age and sex based on betas computed within the HC subpopulation. No other data was corrected for covariates.

##### Model testing

After training and optimization in the CV1 level, the resulting models were applied to the corresponding CV2 fold by preprocessing the best discriminative variables using the learned scaling from the CV1 cycle and determining each testing individual’s label through majority voting across all ensemble models. In other words, in each variable evaluation step in the CV1 level, the SVR algorithm modeled linear relationships between features and labels. Due to the repeated nested cross-validation framework, an ensemble of 100 models (n=10 repetition x k=10 folds) for each CV2 partition (“CV1 ensemble”) was created. Furthermore, due to the 10 repetitions of the CV2 cycle, we were able to establish a final out-of-training label prediction for a given individual by combining all CV1 ensembles into a larger CV2 ensemble, in which the given individual had not served for model training and optimization at the CV1 level. This ensemble generation procedure has been described previously^19^ and is a feature of the model generation and validation process implemented in NeuroMiner.

##### Model significance

To assess statistical significance of the SVR models, we employed permutation testing.^20^ We performed 5000 random permutations of the outcome labels. For each permutation, we retrained the model in the repeated NCV structure using the respective feature subsets obtained from the observed-label analyses. For each permutation, we accumulated the predictions of the random models into a permuted ensemble prediction for each CV2 subject. Thus, we built a null distribution of out-of-training regression performance (MAE) and then calculated the significance of the observed out-of-training MAE as the number of events where the permuted out-of-training MAE was higher or equal to the observed MAE divided by the number of permutations performed. The significance of the model was determined at q=0.05.

The SVR models were applied onto the replication dataset using the same preprocessing pipeline as in the discovery sample. Specifically, the replication data was stratified to the identical CV structure used for model generation and the predictions were pooled across the CV structure to retrieve a cross-validated performance of the SVR model in the replication sample. No further retraining was conducted within the CV structure.

#### Detailed SVR Pipeline

In detail, the machine learning pipeline consisted of the following steps:

##### Repeated nested cross-validation

- CV1 cycle: 10 folds and 10 permutations
- CV2 cycle: 10 folds and 10 permutations

##### Preprocessing Setup

1. Feature-wise Scaling from 0 to 1, zeroing-out completely non-finite features.
2. Pruning of non-informative features with zero variance.
3. Imputation within in matrix block using the median of 7 nearest neighbors (k-nearest-neighbor imputation).
4. Only for EEG as predictor data: attenuating covariate effects (age, sex) via partial correlations. Beta values computed within the HC subpopulation and then used for correction within the entire sample.
5. Feature-wise Scaling from 0 to 1, zeroing-out completely non-finite features.

##### Machine Learning Algorithm Parameter Setup

1. ML model performance criterion: Mean Absolute Error (MAE)
2. Target scaling: [0, 1]
3. Selection and Configuration of ML algorithm

a. LIBSVM 3.1.2 with instance weighting support
b. Regressor type: epsilon-SVR
c. Kernel type: linear
d. Cache size: 500 MB
e. Termination criterion: 0.001
f. Shrinking heuristics: enabled

##### Model Optimization Parameters

1. Slack/Regularization parameters: [0.015625, 0.03125, 0.0625, 0.125, 0.25, 0.5, 1]
2. Eps-SVR parameters: [0.05, 0.075, 0.1, 0.125, 0.15, 0.2]
3. No regularization of model selection
4. Cross-parameter model selection process: Single optimum model (no ensemble)

#### Visualization of predictive pattern elements

We used two computational approaches to visualize the predictive patterns of the SVR models. The detailed mathematical background of these procedures has already been described in the Supplementary Material of Koutsouleris et al.^21^. First, sign-based consistency mapping was employed to measure predictive stability. It is based on an approach proposed by Gómez-Verdejo et al.^22^ toward wrapper-based feature selection strategies. This method assesses how consistently a feature was weighted either positively or negatively across all cross-validated models. In doing so, a *P* value can be computed, measuring whether the distribution of positive and negative weightings of a certain feature across all computed models is significantly different from a random null distribution. The *P* value therefore indicates whether a certain feature received predominantly positive or negative weights above chance level. Thus, sign-based consistency mapping allows us to identify those features, which have been significantly predictive for higher or lower label values. For those features that passed the sign-based consistency threshold and were therefore stable, significant predictors, we calculated the cross-validation ratio (CVR), by computing the grand mean and standard error of all SVR weight vectors across all computed models. The CVR as a measure for the extent of which a pattern was weighted towards one of the labels. It is inspired by the bootstrap ratio commonly used in the Partial Least Squares literature and described in Krishnan et al.^1^. Thus, the CVR indicates how predictive a certain feature was for higher or lower label values. While the direction of the CVR (e.g., positive, or negative) indicates the direction of the label prediction, the absolute value of the CVR indicates how strong the contribution of the feature was for the overall prediction. Combining these two approaches allowed us to identify the most stable and most decisive predictors for our models, therefore giving way to further assessment and interpretation.

#### Software

All analyses were conducted using MATLAB R2022a (MathWorks, Natick, MA, USA). Specifically, we used functions from the MATLAB Statistics and Machine Learning Toolbox as well as the machine learning platform NeuroMiner, release version 1.3, developed by Nikolaos Koutsouleris (https://github.com/neurominer-git/NeuroMiner_1.3)^6,23^

#### MRI Preprocessing Pipeline

The manual of the CAT12 toolbox (https://neuro-jena.github.io/cat12-help/) details the processing steps applied to the structural images. These steps consist of:

1. A 1^st^ denoising step based on Spatially Adaptive Non-Local Means (SANLM) filtering^25^.
2. An Adaptive Maximum A Posteriori (AMAP) segmentation technique, which models local variations of intensity distributions as slowly varying spatial functions and thus achieves a homogeneous segmentation across cortical and subcortical structures.^25^
3. A 2^nd^ denoising step using Markov Random Field approach which incorporates spatial prior information of adjacent voxels into the segmentation estimation generated by AMAP^26^.
4. A Local Adaptive Segmentation (LAS) step, which adjusts the images for white matter (WM) inhomogeneities and varying gray matter (GM) intensities caused by differing iron content in e.g., cortical, and subcortical structures. The LAS step is carried out before the final AMAP segmentation.
5. A Partial Volume Segmentation algorithm that is capable of modeling tissues with intensities between GM and WM, as well as GM and cerebrospinal fluid (CSF) and is applied to the AMAP-generated tissue segments.
6. A high-dimensional DARTEL registration of the image to an MNI-template generated from the MRI data of 555 healthy controls in the IXI database (http://brain-development.org/)

#### MRI Data Quality Assurance

To assess homogeneity of the acquired MRI scans and assure a high standard of MRI data quality, we employed the homogeneity check option of the CAT12 toolbox. As part of the preprocessing, CAT12 calculates several individual quality measures for each MRI scan: NCR (Noise Contrast Ratio), ICR (Inhomogeneity Contrast Ratio) and RES (RMS resolution). These measures are combined into the weighted average image quality rating (IQR). The IQR measure is scaled from 0.5 to 10.5, where 0.5 is a ‘perfect/excellent’ score and 10.5 is deemed ‘unacceptable/failed’. Values around 1 and 2 represent ‘(very) good’ image quality, whereas values of 5 and higher indicate problematic images^27^. The data quality features were entered into the CAT12 “check homogeneity” module along with modulated (m) normalized (w) GM segments (p1). We then calculated the Mahalanobis distance between the mean correlation and weighted overall image quality. Mean correlation quantifies the homogeneity of all selected MRI data used for statistical analysis and is therefore a measure of image quality after pre-processing. The weighted overall image quality combines measurements of noise and spatial resolution of the images before pre-processing. Hence, calculating the Mahalanobis distance between these two measurements estimates image quality both before and after pre-processing. Following this approach, we only included cases with an overall image quality rating (IQR) of “good” to “very good”. This led to the exclusion of one case, which deviated from the rest of the sample by more than two standard deviations. This protocol closely follows the general recommendation as given in the official CAT12-Manual (https://neuro-jena.github.io/cat12-help/). Since MRI data quality is influenced by the individual’s age and often correlates with symptomatology^28,29^ we assumed a potential dimensional impact of data quality in our sample of mentally ill adolescents and young adults, even after passing the data quality assurance protocol. Since machine learning techniques such as sparse partial least squares (SPLS) can potentially detect and be driven by such subtle yet confounding factors, we included IQR as a feature into the analysis

## Notes

### Author Declarations

Ethics committee of the Faculty of Medicine, LMU Munich, Project 17-880, 29.03.2018; project 18-716, 15.10.2020) and at the MPI for Psychiatry (approved by the local ethics committee of the Faculty of Medicine, LMU Munich, project numbers 350-14, 19-310, 20-314,19-678 and 18-393

